# *SPRED1* variants reveal differential impacts on signaling dynamics

**DOI:** 10.64898/2026.07.08.26357239

**Authors:** Juan Báez-Flores, Alessio Coppola, Noemi Ferrito, Ana María Palade, David Hansoe Heredero-Jung, Elena Marcos-Vadillo, Saturnino Ortiz-Madinaveitia, María Isidoro-García, Jesus Lacal, Pablo Prieto-Matos

## Abstract

Germline loss-of-function mutations in *SPRED1* impair its negative regulatory function within the RAS-MAPK signaling pathway, leading to pathway hyperactivation. Clinical sequencing of *SPRED1* frequently identifies variants of uncertain significance (VUS) in patients suspected of Legius syndrome (LS), yet the functional consequences of most variants, specifically their impact on RAS-MAPK regulation, often remain unresolved. Here, we integrate clinical characterization, 3D structural modeling, stable lentiviral expression systems, real-time live cell imaging and multi-pathway signaling profiling to functionally assess a panel of eleven *SPRED1* variants, including two uncharacterized patient-derived alterations, the splice-site mutation c.207+1G>A and the missense substitution c.986A>T (classified as VUS in ClinVar). Unlike c.986A>T, which selectively impairs ERK signaling, the c.207+1G>A variant acts as a broader pathway disruptor, elevating both ERK and p38 phosphorylation, a difference that correlates with its more severe clinical phenotypes. Expanding our analysis across nine additional truncating and missense variants, we uncover reproducible differences in SPRED1 protein abundance and variant-specific signaling dynamics, implicating not only the canonical RAS/ERK axis but also AKT, p38, and p53 pathways. Collectively, these findings reveal genotype-phenotype correlations with differential impacts on signaling dynamics by SPRED1 mutants.

## Introduction

The *SPRED1* gene encodes the Sprouty-related EVH1 domain-containing protein 1 (SPRED1), a key negative regulator of the RAS/MAPK signaling pathway that modulates cellular functions by controlling the spatial and temporal dynamics of MAPK activation. Through its interaction with neurofibromin, SPRED1 constrains RAS activity, thereby contributing to the fine-tuning of cellular proliferation and differentiation. Consequently, *SPRED1* is classified as a central RASopathy-associated gene, as germline disruption of this regulatory axis is a defining feature of RASopathies, a group of developmental disorders caused by germline variants affecting components of the RAS/MAPK signaling pathway^1^.

Heterozygous germline loss-of-function *SPRED1* variants have been described in patients affected by Legius syndrome (LS), an autosomal dominant developmental disorder. LS is primarily defined by the presence of café-au-lait macules (CALMs), axillary and inguinal freckling, macrocephaly, and variable neurodevelopmental features that phenotypically overlap with neurofibromatosis type 1 (NF1)^2–4^. However, the LS phenotype lacks the tumor predisposition observed in NF1, excluding manifestations such as neurofibromas, optic pathway gliomas, or Lisch nodules. This crucial divergence underscores the non-redundant, context-specific role of SPRED1 within the RAS/MAPK network^5^. Additional manifestations reported in LS include mild cognitive impairment, developmental delay, and behavioral phenotypes such as attention deficit/hyperactivity disorder (ADHD) or autism spectrum traits^5^.

The estimated prevalence of LS ranges from 1:46.000 to 1:75.000 based on the frequency of pathogenic *SPRED1* variants identified in NF1 referral cohorts^4,6–9^. To date, more than 170 unrelated individuals carrying germline *SPRED1* variants have been reported^2,4,5,7,10–21^. Beyond LS, *SPRED1* variants have been identified in approximately 2% of tumors according to The Cancer Genome Atlas (TCGA)^12^. Emerging evidence suggests a modestly increased risk for specific malignancies, including childhood leukemia^13^, mucosal melanoma^22^, gastric carcinoma^23^, squamous cell carcinoma^24^, breast cancer^25^, hepatocellular carcinoma^26^, and adult acute myeloid leukemia (AML)^27^. Specifically, in mucosal melanoma, 37% of cases exhibit SPRED1 loss-of-function, frequently occurring in tumors lacking *BRAF*-*RAS* or *NF1* variants, suggesting that SPRED1 inactivation may act as an alternative mechanism of MAPK pathway activation^22^. Consistently, reduced SPRED1 expression correlates with increased ERK phosphorylation and hyperactivation of the RAS/MAPK pathway, supporting a tumor suppressor role for SPRED1^27,28^.

Recent work has further demonstrated that neurofibromin and SPRED1 cooperate through RAS-MAPK-independent mechanisms to modulate MAPK-AKT signaling, revealing that SPRED1 loss phenocopies neurofibromin loss and engages AKT regulatory functions that extend beyond canonical RAS inhibition^29^. Mechanistically, downstream of receptor tyrosine kinases (RTKs) activation, SPRED1 is recruited to the plasma membrane through its interaction with KRAS within BRAF-associated membrane nanodomains^30–34^. Once localized at the cell surface, SPRED1 functions as a molecular scaffold that recruits neurofibromin, a major Ras GTPase-activating protein (RasGAP), thereby promoting the hydrolysis of active RAS-GTP to inactive RAS-GDP and attenuating RAS signaling^3,34^. The assembly and activity of the SPRED1-neurofibromin complex are dynamically regulated by post-translational modifications, including phosphorylation of SPRED1 at serine 105 (S105), which modulates SPRED1-dependent control of RAS signaling^34^. As a consequence of neurofibromin-mediated RAS inactivation, RAF activation is suppressed, including reduced phosphorylation of the regulatory S338 residue, leading to suppression of the MEK/ERK signaling cascade and the subsequent modulation of cell proliferation and differentiation^35^. SPRED1 comprises three distinct functional domains including an N-terminal Enabled/VASP homology-1 (EVH1) domain, a central kit binding domain (KBD), and a C-terminal cysteine-rich Sprouty-related (SPR) domain. Pathogenic *SPRED1* mutations in LS are distributed across all three domains, disrupting distinct molecular mechanisms, including EVH1-mediated neurofibromin recruitment, KBD-dependent signaling modulation, and SPR-driven membrane localization, yet converge on the loss of RAS/MAPK suppression^3,4,12,18,30–43^. In addition to its primary role in MAPK regulation, SPRED1 has been implicated in the modulation of the p38 MAPK pathway, cellular responses to inflammation and stress^44,45^, as well as in functional crosstalk with p53 during cell cycle regulation and the DNA damage response^46^.

To date, *SPRED1* functional studies have primarily relied on transient expression systems or domain-focused analyses^3,18,36,47^. Despite these advances, it remains unclear how variants distributed across distinct SPRED1 domains translate into differential effects on cell signaling. In this study, we aim to overcome this limitation by integrating clinical findings, structural insights, and functional assays in stable cellular models to investigate the impact of *SPRED1* variants associated with both LS and cancer.

## RESULTS

### *SPRED1* variants are uniformly distributed across the entire protein without evidence of mutational hotspots

To assess the distribution and clinical relevance of *SPRED1* variants, we analyzed 846 entries from ClinVar (Supplementary Table S1). Variants were distributed across the entire protein without evidence of mutational hotspots (Fig. 1a). This pattern was maintained when considering pathogenic and likely pathogenic variants only (*n*=112), indicating the absence of domain-specific clustering (Fig. 1b). Normalization of variant frequency by domain length shows that 27% mapped to the EVH1 domain, 23% to the KBD, 29% to the SPR domain, and 22% to interdomain regions, demonstrating a relatively uniform distribution across SPRED1 functional regions. Consistently, no statistically significant association between variant type and domain localization was detected (Fisher’s exact test, p>0.05), supporting a model in which pathogenic variants affect SPRED1 function independently of their positional context.

**Fig. 1.**
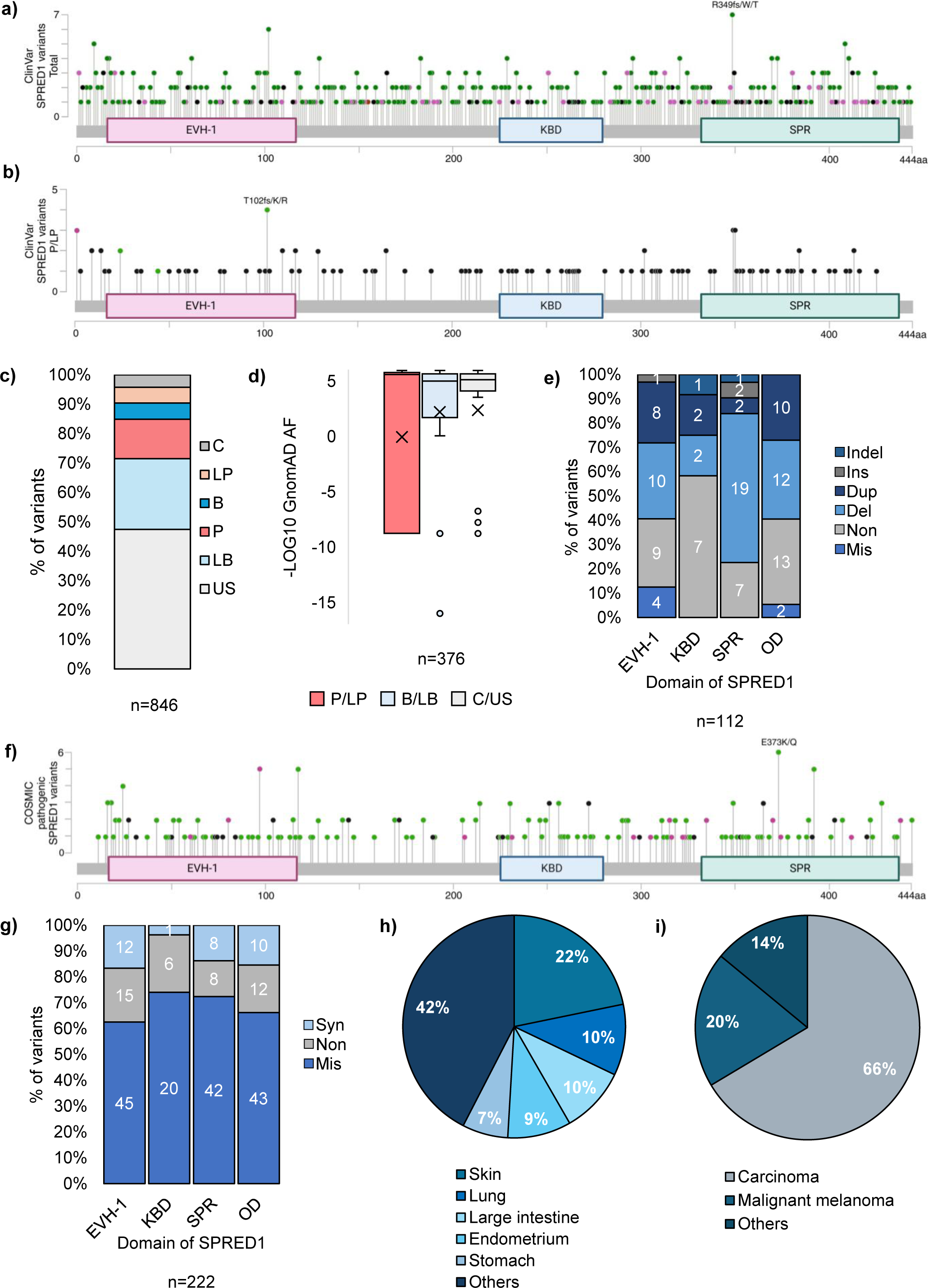
Distribution and classification of germline and somatic *SPRED1* variants. **(a)** Schematic of SPRED1 protein mapping the spatial distribution of 565 ClinVar-deposited *SPRED1* variants out of 846 total entries (retrieved 10/2024), visualized utilizing the cBioPortal mutation mapper tool for cancer genomics (www.cbioportal.org). Lollipops indicators are color-coded by mutation type: truncating (black), missense (green), in-frame (brown), and other alterations (purple). **(b)** One-dimensional linear distribution plot mapping 112 pathogenic/likely pathogenic germline *SPRED1* variants recorded in ClinVar **(c)** Relative frequency distribution of core ClinVar clinical annotations across the 846 variants: P (pathogenic), LP (likely pathogenic), VUS (Variant of uncertain significance), B (benign), LB (likely benign), and C (conflicting interpretation). **(d)** Boxplot quantification of population level-allele frequencies across distinct variant classes extracted from the gnomAD database. **(e)** Spatial compartmentalization of pathogenic *SPRED1* variants categorized by functional domain localization in the protein highlighting occurrences within the EVH1, KBD, SPR, and OD (outside of domains). **(f)** One-dimensional linear plot mapping the distribution of 271 somatic *SPRED1* variants cataloged within the COSMIC cancer database **(g)** Domain-specific structural breakdown of the 222 coding somatic alterations, classifying: missense (*n=*150), nonsense (*n=*41), and synonymous (*n=*31) mutations distributed across SPRED1 domains. **(h)** Organ and tissue-specific profiling of the 271 somatic variants, demonstrating significant enrichment within skin, lung, and large intestine. **(i)** Histological and tumor-type distribution of *SPRED1* somatic variants reported in COSMIC, highlighting a predominant enrichment within carcinomas, with adenocarcinoma representing the most prevalent oncological subtype.

Approximately 15% of ClinVar variants were classified as pathogenic or likely pathogenic, whereas nearly 50% correspond to VUS, highlighting current limitations in variant interpretation and underscoring the critical need for functional validation (Fig. 1c). Consistent with this, analysis of gnomAD frequencies revealed a clear distinction between benign/likely benign and pathogenic/likely pathogenic variants, although benign variants appeared underrepresented, likely reflecting reporting biases in ClinVar (Fig. 1d). Among pathogenic variants, most were associated with LS (*n*=97), with smaller contributions from related phenotypes linked to Noonan-related phenotypes, cardiovascular manifestations, or other genetic disorders (Supplementary Table S1). Truncating variants including deletions, nonsense mutations and duplications predominated, whereas missense variants were comparatively rare. The most common pathogenic variant types include deletions (*n*=43), nonsense substitutions (*n*=36), duplications (*n*=22), missense substitutions (*n*=6), insertions (*n*=3), and insertions/deletions (*n*=2). Their distribution across domains closely mirrored the overall dataset, tracking to the EVH1 (*n*=32), KBD (*n*=12), SPR (*n*=31) and interdomain (*n*=37) regions (Fig. 1e).

To extend this analysis to somatic context, we examined 271 *SPRED1* pathogenic variants annotated in COSMIC (Fig. 1f). Out of these, 222 were located within coding DNA sequences, while the remaining variants mapped upstream of the translation initiation sites (*n*=2), downstream of the translation termination site (*n*=38) or within intronic regions (*n*=9). Most coding alterations were missense substitutions (*n*=150), followed by nonsense (*n*=41) and synonymous variants (*n*=31) (Fig. 1g). Their spatial distribution within SPRED1 shows that 72 variants mapped to the EVH1 domain, 27 to the KBD, 58 to the SPR, and 65 outside defined domains. Like the germline data from ClinVar, somatic variants were evenly distributed across SPRED1 domains, with no evidence of hotspot regions or domain-specific enrichment. Analysis of tissue distribution revealed that these variants are most frequently reported in skin (*n*=59), lung (*n*=28), large intestine (*n*=26), endometrium (*n*=25), and stomach (*n*=18) (Fig. 1h). Among tumor types with histological classification, *SPRED1* variants were significantly enriched in carcinomas (*p<0.05*), accounting for 67% of cases (*n*=180), followed by malignant melanoma (20%, *n*=53) (Fig. 1i). The distribution across carcinoma subtypes includes adenocarcinoma (*n*=68), endometrioid carcinoma (*n*=24) and squamous cell carcinoma (*n*=23). However, no significant association was observed between mutation type and specific carcinoma subtypes. Collectively, these results indicate that *SPRED1* variants are broadly distributed across the protein in both germline and somatic contexts, supporting a model in which pathogenicity arises from global disruption of protein function rather than domain-specific mutational hotspots.

### Interface disruption versus domain destabilization as dual mechanisms of *SPRED1* **missense pathogenicity**

To explore the clinical and functional spectrum of *SPRED1* mutations, we analyzed a panel of eleven variants, including two uncharacterized patient-derived alterations identified at a specialized rare disease diagnostic unit (Fig. 2a). The two *SPRED1* variants identified in unrelated pediatric patients evaluated included the splice-site mutation c.207+1G>A (X69fs*17) and the missense substitution c.986A>T (D329V), neither of which had previously been subjected to detailed molecular or clinical investigation (Fig. 2a). Additionally, we included five previously reported variants associated with LS and found in tumor samples, c.52C>T (R18*), c.184G>A (G62R), c.305C>T (T102M), c.634G>A (V212I) and c.1089A>G(I363M) (Fig. 2a). Also, to assess the functional impact of the predominant truncating mutations in *SPRED1*, we designed four truncated variants including c.478G>T (E160*), c.968C>A (S323*), c.1048G>T (G350*) and c.1318delC (H440Ifs*22) (Fig. 2a). Structural mapping of the missense variants across SPRED1 revealed their distribution within distinct secondary structure elements (Fig. 2b). Variants located in structure regions, including G62R and T102M, within β-sheets and V212I within α-helices, are predicted to disrupt protein folding or local stability. In contrast, variants located in less structured regions, such as the patient derived D329V or the tumor-associated I363M substitution, are predicted to exert more subtle or context-dependent structural effects. Truncating variants showed progressive loss of functional domains, ranging from partial C-terminal deletions to severe truncations retaining only minimal N-terminal sequences (Fig. 2c). Despite their clinical documentation, no functional assays had been performed on these variants prior to this study.

**Fig. 2.**
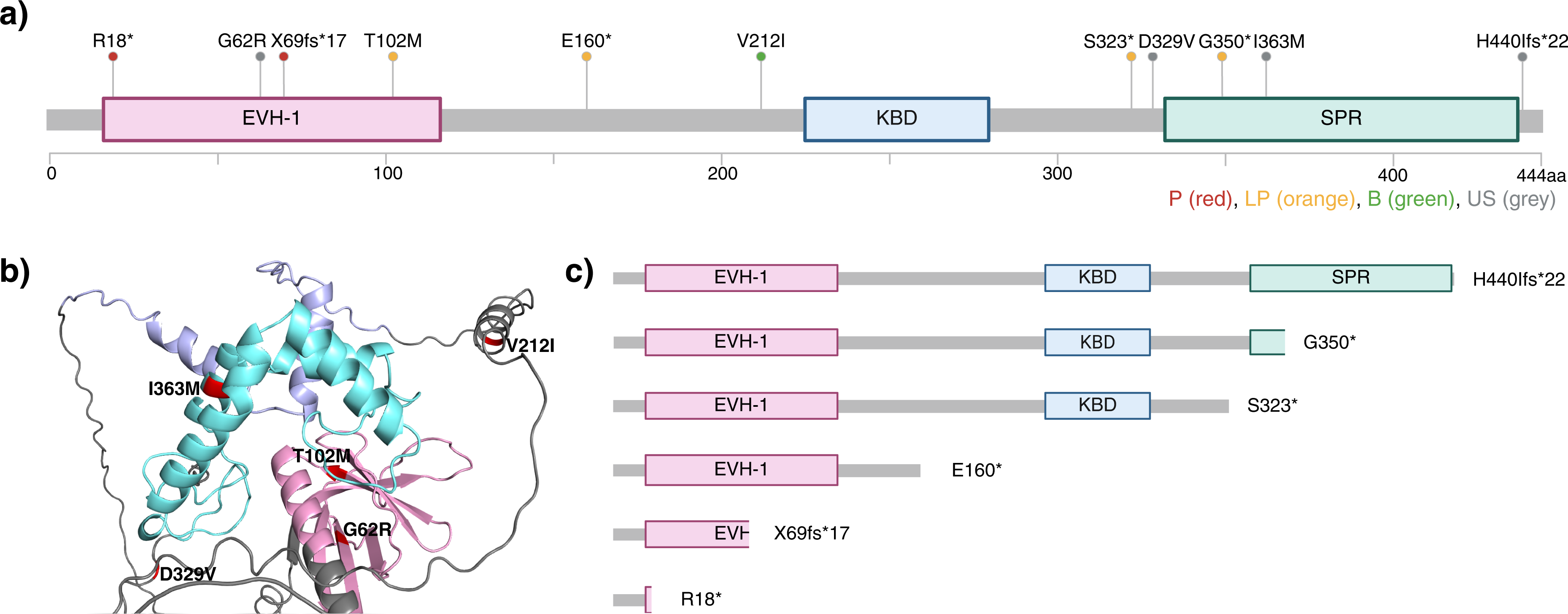
Selection, pathogenic prediction, and three-dimensional structural mapping of *SPRED1* variants panel. **(a)** Comprehensive diagram of the eleven *SPRED1* variants analyzed in this functional study. The panel includes the patient-derived germline variants c.207+1G>A and c.986A>T resulting in X69fs*17 and D329V, identified in unrelated pediatric patients with suspected LS. Five previously documented variants shared between LS cohorts and somatic tumor samples are detailed: c.52C>T, c.184G>A, c.305C>T, c.634G>A and c.1089A>G resulting in R18*, G62R, T102M, V212I, and I363M, respectively. Four generated variants targeting distinct domains are mapped: c.478G>T, c.968C>A, c.1048G>T and c.1318delC, resulting in truncated proteins E160*, S323*, G350*, and H440Ifs*22. Lollipop colors correspond to pathogenic predictions generated via Franklin. **(b)** Three-dimensional structural modeling of the full-length SPRED1 protein mapping the spatial coordinates and side-chain configurations of the investigated missense variants. Protein modules are color-coded by domain boundary definitions: EVH1 domain (pink); KBD (purple) and SPR (mint). **(c)** Linear schematic of truncated SPRED1 protein illustrating the progressive loss of downstream functional domains.

The patient-derived splice-site mutation c.207+1G>A and the truncating variant c.52C>T (R18*) were consistently classified as pathogenic (P) across Franklin, Varsome, and ClinVar (Table 1). In contrast, the patient-derived missense variant c.986A>T (D329V) was classified as uncertain significance (VUS) by all predictors, with no population frequency reported in gnomAD (Table 1). The tumor-associated missense variant c.634G>A (V212I) was consistently classified as benign/likely benign, while c.1089A>G (I363M) and c.1318delC (H440Ifs*22) remained of uncertain significance. Highly conserved residues (G62R, T102M, D329V) showed elevated PhyloP scores (>7.5), whereas the benign V212I variant exhibited a low score (2.12), supporting the correlation between evolutionary conservation and predicted pathogenicity (Table 1).

**Table 1.** *In-silico* pathogenicity predictions and population prevalence of *SPRED1* variants under study. This table summarizes the predicted structural impact, potential pathogenicity and population prevalence of the eleven *SPRED1* variants. Columns detail the specific nucleotide alteration (HGVS cDNA nomenclature), the corresponding amino acid change, and consensus clinical pathogenicity classifications retrieved from Franklin by Genoox, VarSome, and the ClinVar database. Population-level allele frequencies are extracted from the Genome Aggregation Database (gnomAD). and evolutionary conservation scores from PhyloP100. Abbreviations: P, pathogenic; LP, likely pathogenic; VUS, variant of uncertain significance; B, benign; NR, not reported; Con, conflicting. * Premature end of the protein.

| Variant | Protein | Franklin | Varsome | Clinvar |  | GnomAD |  | PhyloP100 scores |
| --- | --- | --- | --- | --- | --- | --- | --- | --- |
|  |  |  |  | Potential pathogenicity | Population prevalence | Allele count | Allele frequency |  |
| c.52C>T | p.R18* | P | P | P | 6 | 1 | 6.46e-7 | na |
| c.184G>A | p.G62R | VUS | P | NR | NR | NR | NR | 9.189 |
| c.207+1G>A | p.X69fs*17 | P | P | LP | 1 | NR | NR | na |
| c.305C>T | p.T102M | LP | P | VUS | 1 | 2 | 1.24e-6 | 7.905 |
| c.478G>T | p.E160* | LP | LP | NR | NR | NR | NR | na |
| c.634G>A | p.V212I | LB | B | Con | 4 | 66 | 4.09e-5 | 2.12 |
| c.968C>A | p.S323* | LP | LP | NR | NR | NR | NR | na |
| c.986A>T | p.D329V | VUS | VUS | NR | NR | NR | NR | 7.56 |
| c.1048G>T | p.G350* | LP | P | NR | NR | NR | NR | na |
| c.1089A>G | p.I363M | VUS | VUS | VUS | 1 | 22 | 1.36e-5 | 0.236 |
| c.1318delC | p.H440Ifs*22 | VUS | NR | NR | NR | NR | NR | na |

To assess the functional relevance of the missense variants, we analyzed their predicted pathogenicity in relation to evolutionary conservation and structural context. The results showed that the EVH1-localized variants G62R and T102M affect highly conserved residues (ConSurf score ≥8) and are consistently predicted as deleterious, whereas V212I and I363M affect poorly conserved positions and show uncertain classifications (Fig. 3a). Mapping ConSurf scores onto SPRED1 structural models confirmed that the EVH1-localized variants G62R and T102M reside within highly conserved clusters, whereas non-EVH1 variants displayed heterogeneous patterns (Fig. 3b). The patient-derived D329V substitution mapped to a highly conserved residue within the linker region, whereas I363M within the SPR domain exhibited intermediate conservation (Fig. 3b). Further analysis with structural modeling revealed that T102 resides within the SPRED1–neurofibromin interaction interface (≤6 Å), directly contacting the GRD domain of neurofibromin, while G62R localizes outside this interface, suggesting distinct pathogenic mechanisms (Fig. 3c–e). These structural predictions provide a biophysical basis consistent with the divergent variant-specific protein expression and signaling profiles characterized in this study. Consistent with these results, thermodynamic calculations revealed that G62R severely destabilizes the protein (ΔΔG_folding = +6.39 ± 2.20 kcal/mol), whereas T102M moderately affects stability (ΔΔG_folding = −1.95 ± 0.01 kcal/mol) but uniquely impairs binding affinity (ΔΔG_binding = −0.58 ± 0.30 kcal/mol), further supporting the distinct pathogenic mechanisms between these two EVH1-localized variants (Supplementary Table S2). These findings indicate that SPRED1 missense pathogenicity correlates strongly with evolutionary conservation yet operates through distinct molecular mechanisms, ranging from global domain destabilization to subtle allosteric effects.

**Fig. 3.**
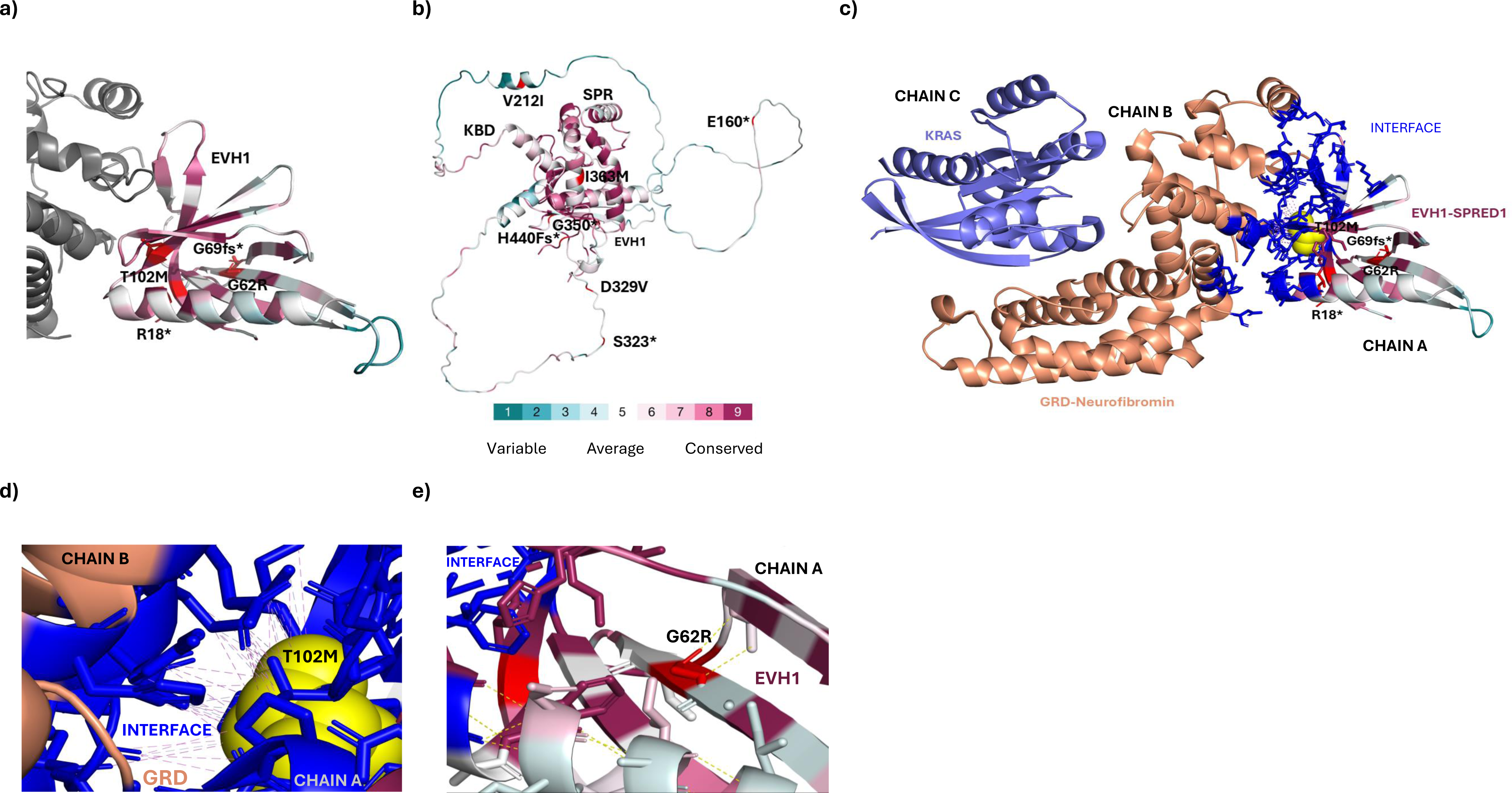
Evolutionary conservation profiles and structural mapping of SPRED1-NF1 interaction interface. **(a)** Mapping of evolutionary conservation scores generated via the ConSurf server onto the crystal structure of the human SPRED1–NF1–KRAS ternary complex (PDB: 6V65) for the N-terminal EVH1 domain. **(b)** Conservation score mapping extended onto the full-length AlphaFold tertiary structure model of human SPRED1, illustrating the spatial topography of non-EVH1 variants within the central linker region and the C-terminal SPR domain. **(c)** Detailed view of the binding interface between the SPRED1 EVH1 domain and the neurofibromin GAP-related domain (PDB: 6V65). Interatomic contacts within an interface threshold distance of ≤6 Å are highlighted to delineate structural positioning. **(d)** Close-up structural analysis of the core SPRED1–neurofibromin interaction interface focusing on the highly conserved T102 residue, mapping the specific residue coordination networks and direct polar contacts (≤4 Å) that stabilize the wild-type complex architecture, alongside the predicted impact of the c.305C>T (T102M) substitution. **(e)** Structural localization of the c.184G>A (G62R) missense variant; the residue is positioned distal to the neurofibromin binding interface, indicating that its pathogenic functional defect is driven by local domain destabilization or folding integrity disruption rather than direct steric hindrance of neurofibromin recruitment.

### Clinical spectrum of *SPRED1* c.207+1G>A and c.986A>T variants reveals divergent clinical expressivity and incomplete penetrance in pediatric patients with LS

We identified two *SPRED1* variants in unrelated pediatric patients with clinical features suggestive of neurofibromatosislrelated disorders (Figure 4). These variants correspond to the splice-site mutation c.207+1G>A (X69fs*17) and the missense substitution c.986A>T (D329V), neither of which had previously been subjected to detailed molecular or clinical investigation. Segregation analysis via parental sequencing confirmed autosomal dominant inheritance for both, the c.207+1G>A in Cases I and Ia, and c.986A>T in Cases II and IIb (Table 2). A pediatric patient (Case I) carrying the heterozygous c.207+1G>A *SPRED1* variant presented with multiple CALMs. Clinical evaluations revealed macrocephaly and mild short stature, and neurodevelopmental features consistent with LS (Table 2). The proband also exhibited Noonan-like facial features, specifically hypertelorism, low-set ears with a thickened helix, curly hair and the CALMs (clinical images available upon request). Consistent with autosomal dominant inheritance, Case Ia carried the same variant and displayed a concordant LS phenotype, including multiple CALMs and Noonan-like facial features (Table 2). Both individuals lacked neurofibromas, Lisch nodules, osseous/vascular lesions, and other NF1-associated complications, consistent with a classical LS phenotype. Morphological assessment of the extremities of Case I and Ia showed no structural hand abnormalities, excluding overlapping digital malformations often observed in other RASopathies. Both individuals presented with normal brain magnetic resonance imaging (MRI) findings (Table 2).

**Fig. 4.**
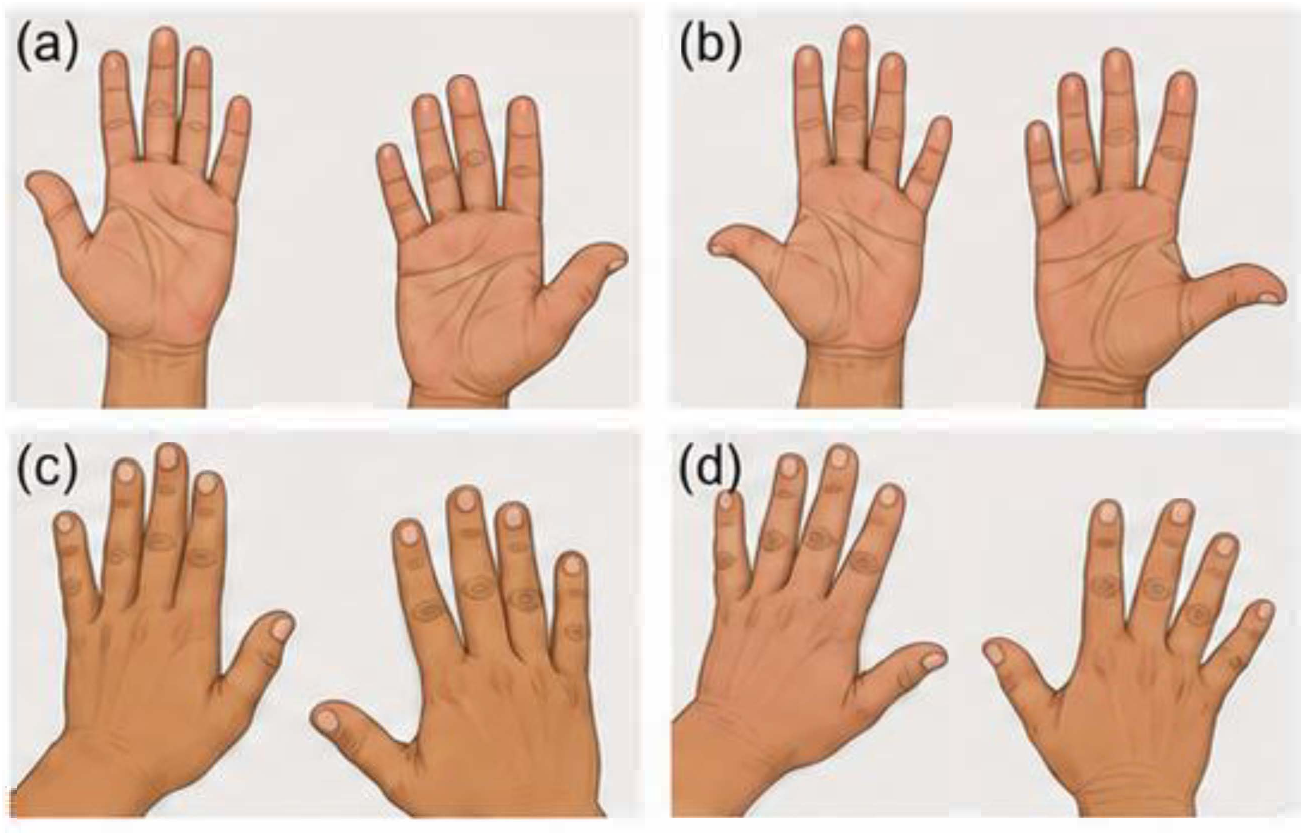
Clinical phenotypes images of pediatric patients diagnosed with LS. Clinical images are not included in this preprint to protect patient privacy. Readers may request access to clinical documentation by contacting the corresponding author at

**Table 2.**
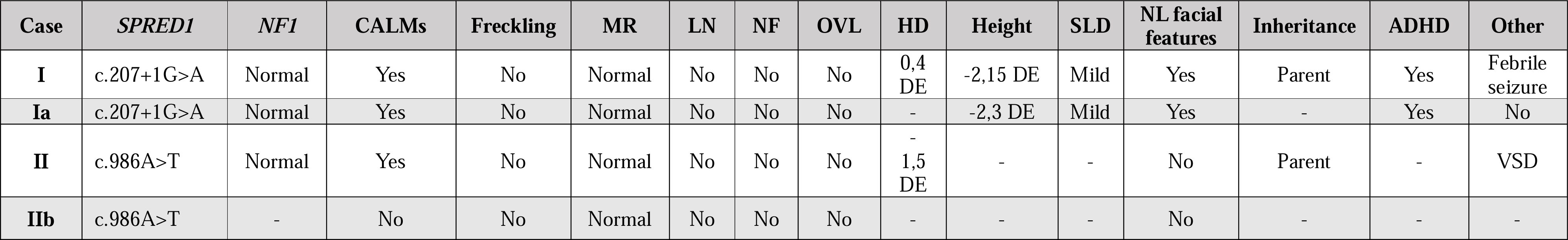
Clinical and genetic features of pediatric probands and family carriers with suspected neurofibromatosis-related phenotypes. This table summarizes the genetic variants, inheritance pattern, and principal clinical features evaluated across the four cases. Detailed clinical data are available upon request to the corresponding author at

A second pediatric patient in their early teens (Case II) carrying the c.986A>T *SPRED1* variant, exhibited multiple CALMs and macrocephaly consistent with LS (Table 2). No additional LS-associated features were observed, and MRI findings were normal (Table 2). The condition was inherited from Case IIb, who showed no clinical features associated with LS (Table 2). Both individuals lacked neurofibromas, Lisch nodules, osseous/vascular lesions, and other NF1-associated complications, consistent with a classical LS phenotype. This inheritance from an asymptomatic carrier Case IIb indicates potential incomplete penetrance of the c.986A>T variant and suggests a milder functional impact. These findings highlight the intrafamilial variability in clinical expressivity associated with *SPRED1* variants in LS patients, emphasizing the importance of genetic and phenotypic characterization for accurate diagnosis.

### Genetic variation differentially affects SPRED1 protein expression levels in HEK293T cells

Despite the growing number of *SPRED1* variants identified through clinical sequencing, the functional impact of most remains unexplored. In this regard, WT *SPRED1* and eleven variants were stably expressed in HEK293T cells. Consistent with previous reports of low basal *SPRED1* expression in HEK293T cells, endogenous SPRED1 was barely detectable in parental and MOCK-transduced samples (Fig. 5a-b). As expected, all of the generated cell lines, but c.52C>T and c.207+1G>A, consistently showed elevated SPRED1 protein levels, with reproducible expression levels across independent biological replicates (Fig. 5a-b), suggesting that the observed differences in protein abundance reflect consistent properties of each variant-expressing cell line rather than stochastic integration effects. Truncated proteins generated by the c.52C>T and patient-derived c.207+1G>A variants were not detectable by WB due to loss of the specific antibody epitope located at position 134-240 of the SPRED1 protein. However, RT-qPCR confirmed high levels of the corresponding mRNAs, indicating that these variants were successfully transduced (Supplementary Fig. S1a). Interestingly, expression of the c.52C>T variant appeared to correlate with a slight increase in a band corresponding to the molecular weight of endogenous SPRED1 (Fig. 5a; Supplementary Fig. S1b). This result suggests that this variant may interfere with global proteostasis, potentially affecting the turnover of endogenous protein, a phenomenon previously described for other truncated or misfolded proteins where aberrant protein products alter ubiquitin-proteasome dynamics or promote compensatory upregulation of the endogenous protein.

**Fig. 5.**
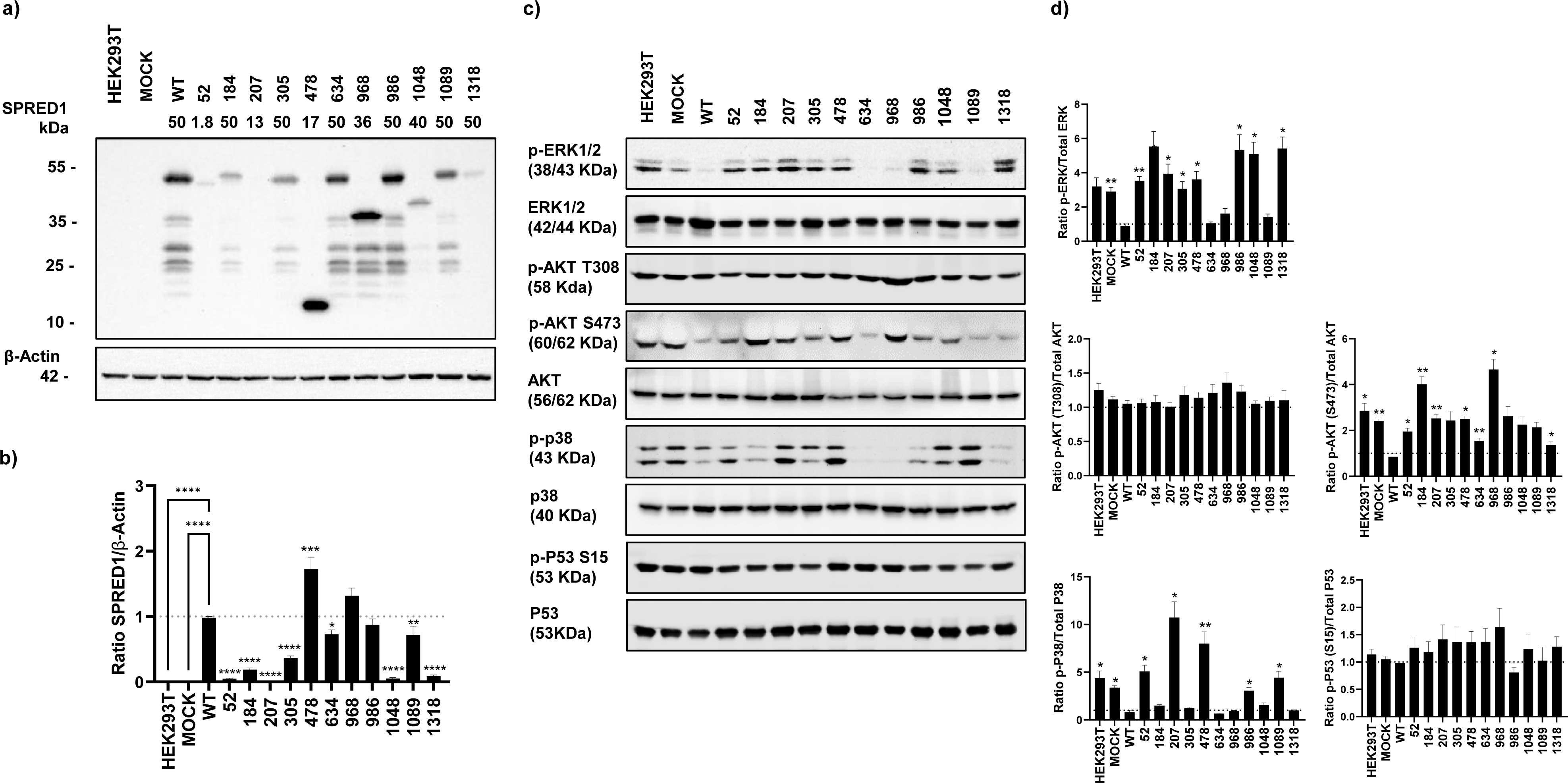
Characterization of steady-state protein expression and multi-pathway signaling cascade modulation across the *SPRED1* variant panel. **(a)** Protein expression analysis by Western blot in HEK293T cells. WT *SPRED1* or variant protein levels were normalized to β-Actin as a loading control. **(b)** Quantification of the relative SPRED1 protein abundance. Data are expressed as mean ± SEM (*n* = 5 independent biological replicates). Statistical significance was determined by a one-way ANOVA followed by Dunnett’s post-hoc test for multiple comparisons against the WT reference control. Significant differences in protein expression levels are defined as ****P<0.0001, ***P<0.001, **P<0.01, and *P<0.05. **(c)** Western blot analysis of key signaling pathways in HEK293T cells expressing WT *SPRED1* or variants. Total and activation levels of the biomarkers ERK, AKT, p38 and p53 were assayed. WT SPRED1 reduces ERK and AKT phosphorylation, while variants exhibit differential effects on pathway regulation. **(d)** Analysis and relative ratio quantification of active phosphorylated cascades to their respective total protein baselines (Ratio p-ERK/Total ERK, Ratio p-AKT Thr308/Total AKT, Ratio p-AKT Ser473/Total AKT, Ratio p-p38/Total p38, and Ratio p-p53 Ser15/Total p53), with the WT control group set as the reference value of 1.0. Data are presented as mean ± SEM from independent biological replicates (ERK and p53, *n* = 5; AKT Thr308, AKT Ser473 and p38, *n* = 4). Statistical significance was assessed using repeated-measures one-way ANOVA followed by Dunnett’s multiple-comparison test against the WT reference control. Significant differences in protein expression levels are defined as ****P<0.0001, ***P<0.001, **P<0.01, and *P<0.05.

The WB results also show a consistent pattern of lower molecular weight bands (ranging from ∼12 to 40 kDa) across SPRED1-overexpressing cell lines, compatible with recurrent proteolytic processing at several positions along the protein (Fig. 5a). This fragmentation pattern was reproducible across variants, suggestive of systematic proteolytic processing at multiple sites. While the precise cleavage coordinates remain to be determined, the lack of a clear consensus proteolytic motif suggests that this processing may be driven by structural accessibility rather than primary sequence specificity. Uniform β-actin levels across all samples confirmed equal protein loading, ensuring that the observed differences in SPRED1 fragment intensity are variant-specific (Fig. 5a).

### Variant-specific disruption of SPRED1-mediated signaling pathways

While SPRED1 is a well-established negative regulator of the RAS/MAPK pathway, its impact on broader signaling networks remain poorly characterized. Given that RAS activates multiple downstream effectors, including the PI3K/AKT, p38 MAPK, and p53 pathways, we examined these biomarkers to determine how *SPRED1* variants differentially impact broader signaling networks and whether these molecular signatures correlate with clinical variability (Fig. 5c).

As expected, overexpression of WT SPRED1 significantly reduced ERK phosphorylation compared to control conditions, consistent with its established role as a negative regulator of the RAS/MAPK pathway (Fig. 5c). Quantitatively, using the WT-expressing cell line as the reference control for this experiment (set at 1.00), phospho-ERK levels in MOCK and parental HEK293T cells exhibited an approximate 3.3-fold and 3.6-fold increase relative to WT values, respectively, confirming a robust decrease in ERK activation upon WT overexpression (Fig. 5c-d). Among the eleven variants tested, c.634G>A, c.968C>A, and c.1089A>G, suppressed ERK phosphorylation to levels comparable to WT SPRED1 (mean values ∼1.05, ∼1.62, and ∼1.40, respectively), indicating partial preservation of inhibitory function. In marked contrast, the remaining eight variants failed to efficiently suppress ERK phosphorylation. Four variants (c.207+1G>A, c.986A>T, c.1048G>T and c.1318delC) exhibited the highest phospho-ERK levels, reaching mean phospho-ERK values of ∼3.93, ∼5.34, ∼5.09, and ∼5.41, respectively, which represent a significant 4- to 5-fold increase compared to WT (adjusted p < 0.05) (Fig. 5c-d). Importantly, no correlation was observed between SPRED1 protein abundance and ERK phosphorylation status. This dissociation was evident in both directions, c.986A>T maintained near-WT SPRED1 protein levels (0.87-fold, ns) yet showed robust ERK hyperactivation (5.34-fold, p=0.043), while c.1048G>T and c.1318delC showed low SPRED1 expression yet failed to elevate AKT, p38, or proliferation despite strong ERK activation. Conversely, c.634G>A maintained both WT-like SPRED1 expression and WT-like ERK suppression yet showed significantly reduced proliferative expansion (0.84-fold, p=0.0007), indicating that SPRED1-independent mechanisms may contribute to growth regulation in specific variant contexts. Furthermore, c.478G>T and c.968C>A showed elevated SPRED1 levels (1.72-fold and 1.32-fold above WT, respectively) yet displayed impaired signaling regulation across multiple pathways, confirming that protein accumulation does not compensate for functional loss in these truncating variants. These results indicate that most *SPRED1* variants fail to suppress ERK phosphorylation, leading to sustained MAPK pathway activation. Total ERK levels remained consistent across all samples, confirming that the observed differences were specific to phosphorylation status rather than changes in protein expression.

Beyond canonical MAPK signaling, active RAS can engage the PI3K/AKT cascade, a parallel axis central to cell survival, growth, and metabolism. Although SPRED1 is not classically defined as an AKT regulator, the intricate crosstalk between downstream RAS effectors raises the possibility that SPRED1 loss-of-function variants may indirectly influence AKT signaling kinetics. Given that AKT hyperactivation is frequently associated with developmental abnormalities and overgrowth phenotypes, evaluating AKT phosphorylation at Ser473 and Thr308 across our SPRED1 variant-expressing cells could provide insights into how signaling diversion might contribute to patient-specific phenotypes, such as macrocephaly or ventricular septal defects. Our results show that total AKT levels remained consistent across all experimental conditions, as did phospho-AKT levels at Thr308 residue (Fig. 5c), suggesting that the proximal site primarily phosphorylated by PDK1 downstream of PI3K activation, is not substantially affected by SPRED1 status. In contrast, overexpression of WT SPRED1 was associated with a reduction in phosphorylation at Ser473, a residue targeted by mTORC2 and commonly used as a marker of full AKT activation (Fig. 5c-d). This observation suggests that SPRED1 may influence AKT signaling. Among the variants analyzed, eight (c.52C>T, c.184G>A, c.207+1G>A, c.305C>T, c.478G>T, c.968C>A, c.986A>T, and c.1048G>T) failed to suppress Ser473 phosphorylation to the same extent as WT SPRED1, indicating an impaired negative regulation of AKT signaling. In contrast, variants c.634G>A, c.1089A>G and c.1318delC maintained AKT phosphorylation levels comparable to WT SPRED1 (Fig. 5c-d). Two variants, c.184G>A and c.968C>A, exhibited the strongest effects, reaching approximately 4-5-fold higher phospho-AKT (Ser473) levels relative to the WT control (adjusted p < 0.05), indicating a pronounced alteration in AKT signaling dynamics (Fig. 5c-d).

The p38 MAPK is a well-characterized stress-responsive kinase that can be activated downstream of RAS through parallel pathways, including MKK3 and MKK6 kinases. While p38 signaling outputs are highly context-dependent, their dysregulation has been associated with inflammatory responses, neurodevelopmental alterations, and cardiac defects; clinical features that are variably present in patients with LS. In this context, assessing p38 phosphorylation across our *SPRED1* variant-expressing cells may provide insights into whether specific mutations influence alternative MAPK branches, potentially explaining phenotypic variability that cannot be accounted for by ERK hyperactivation alone. The results show that overexpression of WT SPRED1 was associated with reduced p38 phosphorylation compared to control conditions, indicating that its homeostatic regulatory role may extend beyond ERK to stress-activated MAPK pathways. Whereas total p38 levels remained stable across all conditions (Fig. 5c). Several variants, including c.184G>A, c.305C>T, c.634G>A, c.968C>A, and c.1318delC, maintained phospho-p38 levels comparable to WT, suggesting preserved regulatory capacity along this branch. Interestingly, a subset of variants exhibited increased p38 activation (Fig. 5c-d), including the patient-derived-splice-site variant c.207+1G>A (∼13-fold), which showed the strongest effect. This variant was followed by c.478G>T with a ∼9.9-fold increase, c.986A>T (3.06-fold above WT, p=0.034) and the variant c.1089A>G with a ∼5.5-fold (Fig. 5c-d). These differences were statistically significant (repeated-measures ANOVA, p = 0.0047), with post hoc Dunnett’s test confirming significant increases for c.207+1G>A, c.478G>T, and the second patient variant, c.986A>T relative to WT (Fig. 5d). Importantly, c.986A>T showed moderate p38 elevation that was quantitatively and mechanistically distinct from the strong p38 activation observed in c.207+1G>A and c.478G>T, suggesting a different degree of p38 pathway engagement. Rather than indicating a direct inhibitory mechanism, this pattern is highly consistent with a redistribution of mitogenic signaling across parallel MAPK branches as a compensatory response to impaired proximal ERK control.

Although SPRED1 does not directly interact with p53, sustained or aberrant signaling from RAS/MAPK or PI3K/AKT axes can influence p53 stability and transcriptional activity through MDM2-dependent mechanisms. Examining p53 expression and phosphorylation (Ser15) levels in SPRED1 variant-expressing cells could reveal whether certain loss-of-function mutations impose chronic proliferative or genotoxic stress thereby engaging p53 as a compensatory tumor-suppressive response. However, our results show that WT SPRED1 expression did not activate or suppress baseline p53 signaling, as both total and phospho-p53 (Ser15) levels remained fully comparable to control conditions. Similarly, none of the eleven variants tested showed statistically significant differences relative to WT or control conditions (Fig. 5c-d).

### *SPRED1* variants differentially modulate cell proliferation in a variant-specific manner

Because the signaling analyses identified differential effects on ERK, AKT and p38 activation, we next examined whether these molecular signatures were associated with alterations in proliferative behavior. To do so, cell proliferation was monitored in real-time using the IncuCyte® live-cell imaging system coupled with AI-based nuclei object detection. Cell growth was quantified as the fold-change in nuclei object counts relative to the initial time point (t = 0h) and kinetic profiles were recorded across a 72-h window to capture both exponential growth dynamics and potential late-phase proliferative changes (Fig. 6a). One-way ANOVA revealed a significant overall effect of SPRED1 variation on cell proliferation (F = 70.73, p < 0.0001, R^2^ = 0.9725). Variants associated with the strongest disruption of SPRED1-mediated signaling generally exhibited reduced proliferative expansion relative to WT SPRED1. This group included c.52C>T, c.184G>A, c.207+1G>A, c.305C>T, c.478G>T and c.634G>A, all of which showed significantly lower cell numbers at 48 h compared with WT cells (adjusted p < 0.002 to p < 0.0001) (Fig. 6b). In contrast, variants c.1048G>T, c.1089A>G and c.1318delC displayed proliferation profiles comparable to WT SPRED1 (p > 0.05), whereas the patient-derived variant c.986A>T exhibited significantly increased proliferative expansion (adjusted p < 0.0001) (Fig. 6b). Interestingly, kinetic analysis revealed that variants differed not only in the magnitude but also in the temporal pattern of cell expansion. c.478G>T displayed the most pronounced progressive deceleration of any variant, with interval growth rates declining from 65% per 12-hour period during early expansion (12–24h) to only 10% in the final interval (48–60h), the lowest rate observed across all conditions. This kinetic pattern is consistent with the sustained strong p38 hyperactivation observed for this variant (7.99-fold above WT, p=0.047), as p38 MAPK is a well-established inhibitor of cell cycle progression. In contrast, c.986A>T maintained high and sustained growth rates throughout the 72-hour window, consistent with its unique combination of ERK hyperactivation and enhanced proliferative capacity. A distinct kinetic pattern was observed for c.968C>A, which displayed an unusually high interval growth rate of a 2.56-fold proliferative burst between 36 and 48 hours the largest single-interval expansion observed across the entire variant panel, followed by a sharp decline of −14% in the subsequent interval (48–60h), suggestive of a transient hyperproliferative episode followed by proliferative collapse. This pattern may reflect a cycle of growth overactivation and subsequent cytostatic response, potentially linked to the exceptionally elevated AKT Ser473 phosphorylation observed for this variant (4.66-fold above WT, p=0.016).

**Fig. 6.**
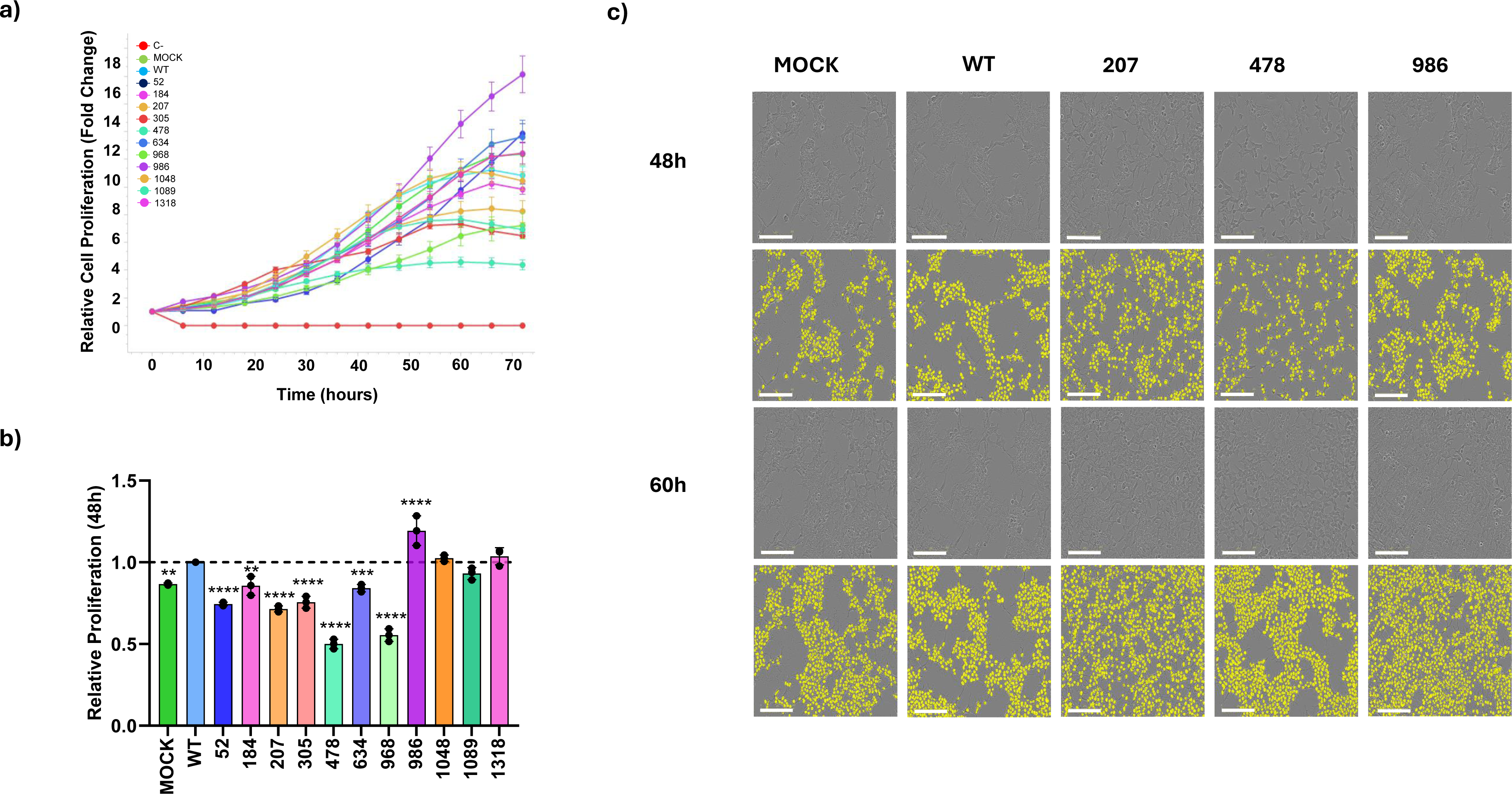
*SPRED1* variants differentially modulate cell proliferation in a variant-specific manner. **(a)** Real-time cell proliferation kinetics of HEK293T cells expressing WT or variant SPRED1 constructs monitored over 72 hours utilizing the IncuCyte® live-cell imaging system. Growth curves represent the automated AI-Based nuclei object count normalized to the initial time point (t = 0 h), plotted as mean ± SEM (*n* = 3 independent biological replicates). **(b)** Quantitative analysis of relative cell proliferation at the 48-hour peak exponential phase derived from the curves in panel (a), expressed as fold-change relative to WT. Statistical validation was executed via repeated-measures one-way ANOVA followed by Dunnett’s post-hoc test (****P<0.0001, ***P<0.001, **P<0.01, *P<0.05 vs. WT). Data are expressed as fold-change relative to WT. Statistical comparisons identify both significantly reduced and significantly increased proliferative expansion across the variant panel. **(c)** Representative phase-contrast images and corresponding overlaid AI-based nuclei segmentation masks (yellow) captured at the 48-h and 60-h analytical intervals across selected key conditions (MOCK, WT, and the clinical variants c.207+1G>A, c.478G>T, and c.986A>T). Scale bar: 200 μm.

Long-term population fitness assessed by Area Under the Curve (AUC) analysis across the full 72-hour kinetic profile confirmed that proliferative differences observed at 48h reflect sustained growth trajectories rather than transient responses (Supplementary Fig. S2a). The c.478G>T variant showed the lowest normalized AUC in the variant panel (AUC ratio = 0.584 relative to WT), fully consistent with its 48h proliferation value (0.469 relative to WT), confirming that the progressive kinetic deceleration accumulates into substantially reduced overall proliferative output. Variant c.634G>A represents a distinct case in which significantly reduced proliferative expansion (0.84-fold, p=0.0007) occurred in the complete absence of detectable signaling alterations across all biomarkers examined, ERK, AKT Ser473, p38, and p53 were all statistically indistinguishable from WT. This observation indicates that SPRED1 may influence cell growth through mechanisms that operate independently of the canonical signaling nodes captured by the current biomarker panel, potentially involving cytoskeletal regulation, cell adhesion dynamics, or alternative downstream effectors not assessed in this study.

Overall, proliferation patterns did not consistently mirror the behavior of any individual signaling biomarker. Variants c.207+1G>A, c.1048G>T and c.1318delC all exhibited strongly elevated phospho-ERK levels but failed to show increased proliferation. In contrast, the patient-derived variant c.986A>T uniquely combined robust ERK hyperactivation with enhanced proliferative expansion, while c.968C>A showed near-WT ERK levels yet markedly reduced proliferation driven by its distinct kinetic profile. To evaluate potential relationships between signaling outputs and proliferative behavior, Pearson correlation analyses confirmed the absence of significant linear associations between proliferation at 48h and any individual signaling biomarker, including phospho-ERK (r=0.31, 95% CI −0.29 to 0.74, p=0.30), phospho-AKT Ser473 (r=−0.44, 95% CI −0.80 to 0.14, p=0.13), and phospho-p38 (r=−0.41, 95% CI −0.78 to 0.18, p=0.16) (Supplementary Fig. S2b). Phospho-AKT Thr308 and phospho-p53 Ser15 were excluded from correlation analyses as neither showed statistically significant variation across the variant panel. The absence of significant correlations indicates that cellular proliferative behavior emerges from the integrated activity of multiple signaling pathways rather than from alterations in any single downstream node.

Unsupervised hierarchical clustering segregated variants into functionally coherent modules driven by combined multi-pathway signatures rather than single-pathway activation states (Supplementary Fig. S2c). Variants characterized by strong p38 activation, particularly c.207+1G>A and c.478G>T, clustered within a low-proliferation module, consistent with the established anti-proliferative roles of sustained p38 activation. The c.986A>T grouped with variants displaying elevated ERK activity but limited p38 engagement, consistent with its enhanced proliferative capacity. Representative phase-contrast images and AI-based segmentation masks confirmed the observed differences in cell density across experimental conditions (Fig. 6c). These results demonstrate that *SPRED1* variants differentially modulate proliferative behavior through variant-specific mechanisms, and that cellular growth responses reflect the integrated output of multiple signaling pathways rather than any single effector node.

## Discussion

Our integrative structural, biochemical, and cellular analysis of *SPRED1* variants reveals that pathogenicity operates through variant-specific mechanisms extending beyond canonical RAS/ERK dysregulation, producing divergent signaling outputs that correlate with clinical heterogeneity. The EVH1 domain emerged as the most functionally constrained region, consistent with its essential role in neurofibromin recruitment^31,34^. Within this domain, our analyses identified two mechanistically distinct classes of missense variants, those such as T102M that localize directly within the SPRED1–neurofibromin interaction interface, and those such as G62R that predominantly affect domain stability and folding without directly disrupting the binding interface. Variant T102M preserved near-WT neurofibromin binding affinity, indicating that the biochemical properties of the substituted residue critically influence functional outcome. This contrasts with the previously reported T102R substitution, which completely abolished neurofibromin binding^34^, and demonstrates that not all interface residues contribute equally to complex stability. T102M may alter local conformational dynamics or regulatory interactions at the EVH1 interface, representing an intermediate loss-of-function phenotype. These observations reinforce the concept that SPRED1 pathogenicity exists along a functional continuum, ranging from complete loss-of-function variants to mutations that preserve partial molecular activity but nevertheless alter network-level signaling outputs and cellular behavior.

Several lines of evidence support a variant-specific interpretation in which protein levels were reproducible across replicates, while full-length variants expressed variably, and no correlation was observed between SPRED1 abundance and ERK phosphorylation, indicating that functional outcomes are independent of expression levels. As expected, disruption of SPRED1 activity resulted in sustained ERK activation in most variants, confirming the central role of SPRED1 as a negative regulator of the RAS/MAPK pathway. However, the signaling consequences of SPRED1 dysfunction extended considerably beyond the canonical pathway. Although SPRED1 is not traditionally considered a direct regulator of PI3K/AKT signaling, eight of the eleven variants analyzed failed to suppress AKT phosphorylation at Ser473 while leaving Thr308 phosphorylation largely unaffected. Interestingly, a bidirectional relationship between SPRED1 and AKT has been suggested in other contexts. In FLT3-ITD-driven AML, AKT-mediated serine phosphorylation of SPRED1 stabilizes and activates it, diverting its function toward miRNA biogenesis regulation rather than canonical RAS/MAPK suppression^48^. This suggests that the SPRED1–AKT axis may operate through distinct, context-dependent mechanisms depending on the cellular environment and the nature of SPRED1 dysfunction. Consistent with our findings, a recent study demonstrated that SPRED1 loss cooperates with neurofibromin loss to modulate MAPK-AKT signaling through mechanisms that are at least partially independent of RAS activation, providing a mechanistic framework for the selective AKT Ser473 dysregulation observed across our variant panel^29^. Similarly, several variants, most notably c.207+1G>A, induced robust p38 hyperactivation, consistent with redistribution of mitogenic signaling across parallel MAPK branches following impaired proximal ERK control^49,50^, rather than direct pathway activation.

The proliferation studies revealed that variant-dependent cellular growth responses could not be predicted by any individual signaling biomarker. Variants with comparable ERK hyperactivation frequently displayed different proliferative outcomes. Variants characterized by strong p38 activation, particularly c.207+1G>A and c.478G>T, clustered within a low-proliferation module, consistent with the well-established anti-proliferative roles of sustained p38 activation. The case of c.634G>A is particularly instructive, despite showing no significant alteration in ERK, AKT Ser473, p38, or p53, this variant produced a statistically significant reduction in cell proliferation (p=0.0007), indicating that SPRED1 may influence cell growth through mechanisms that operate independently of the canonical signaling nodes captured by the current biomarker panel, potentially involving cytoskeletal regulation, cell adhesion, or alternative downstream effectors.

Based on their combined profiles, the eleven variants grouped into three functional classes providing a practical framework for interpreting variant-specific clinical heterogeneity: c.634G>A, c.968C>A and c.1089A>G, which preserve ERK suppression and near-WT proliferation despite variable protein levels; (ii) c.207+1G>A, c.478G>T and c.52C>T, characterized by multi-pathway dysregulation including p38 hyperactivation, and markedly reduced proliferation; and (iii) c.986A>T, c.1048G>T, c.1318delC, which display selective ERK hyperactivation with preserved protein levels but diverge in proliferation, with c.986A>T uniquely enhancing growth.

The molecular findings are directly reflected in the clinical presentations of the two patient-derived variants. The severe and multi-pathway signaling alterations induced by c.207+1G>A are consistent with the classical LS phenotype, and the clinical manifestations observed in Case I and Case Ia. Conversely, the subtler and more selective functional behavior of c.986A>T, combined with its inheritance from a fully asymptomatic carrier (Case IIb), is consistent with incomplete penetrance and suggests that additional genetic, epigenetic, or environmental modifiers contribute to disease expression. These findings align with broader intrafamilial variability documented across RASopathies, where no clear LS genotype–phenotype correlations have been established and even pathogenic *SPRED1* variants can present without café-au-lait macules^18,51^. Analogous patterns in *PTPN11*-related NS, where up to 50% of carrier parents are clinically silent^52,53^, emphasize that the same variant can produce outcomes ranging from full expression to non-penetrance, reinforcing that functional characterization, as performed here, is essential for assessing pathogenicity independently of clinical presentation.

Collectively, our findings demonstrate that quantitative multi-pathway biomarker profiling provides a powerful complementary tool for understanding the molecular mechanism behind LS, supporting the functional interpretation of VUS and improving genotype-phenotype correlations in LS. Future validation in disease-relevant models, such as recently established LS iPSC lines^54^, will enable more precise dissection of variant-specific effects on SPRED1 homeostasis and signaling.

## Materials and Methods

### Patients and SPRED1 data

This study investigated 11 distinct *SPRED1* variants (mRNA NM_152594.3), including two clinical alterations (c.207+1G>A and c.986A>T) identified in unrelated pediatric patients presenting with clinical features suggestive of LS. Patients were evaluated by the clinical team, including experts in rare diseases and pediatric neurologists. Phenotypic data collection was systematically guided by an anthropometric, neurological, and dermatological questionnaire designed according to the current clinical diagnostic criteria for LS.

The variant panel was expanded to include five previously reported variants (c.52C>T, c.184G>A, c.305C>T, c.634G>A, c.1089A>G) associated with LS and documented in somatic tumor registries^15,55^. These variants were obtained from IntOGen and LOVD databases. Python (version 3.9.0) was used for variant sorting and comparison^56^. Additionally, four site-directed truncated variants (c.478G>T, c.968C>A, c.1048G>T and c.1318delC) were engineered to systematically characterize the functional consequences associated with premature protein truncation.

Clinical photographs of the probands and their family members were obtained during routine phenotypic evaluation by the attending clinical team, using conventional digital photography to document dermatological findings (café-au-lait macules, axillary and inguinal freckling), morphological features (upper and lower extremities and facial dysmorphology), and neurodevelopmental assessment in accordance with the current diagnostic criteria for Legius syndrome. Clinical images have been omitted from this preprint to protect patient privacy in accordance with medRxiv policies. Original clinical documentation is retained in the authors’ records. Readers may contact the corresponding author to request access.

### DNA Isolation from patients and NGS

Peripheral whole blood samples were collected from the probands and their parents in EDTA-coated tubes. Genomic DNA (gDNA) extraction and purification were performed using a MagNA Pure Compact automated system (Roche Diagnostics) according to the manufacturer’s instructions. DNA concentration, quality, and purity ratios (A_260_/A_280_) were assessed with a Biophotometer spectrophotometer (Eppendorf). Next-Generation Whole Exome Sequencing (WES) was performed on the NextSeq 550 platform (Illumina®). Genomic libraries were prepared using TruSeq technology (Illumina®), followed by tagmentation and exome target enrichment with the xGen™ Exome Research Panel (Integrated DNA Technologies, IDT), covering exons and adjacent flanking intronic consensus regions of protein-coding genes. Sequencing reads were aligned against the GRCh37/hg19 human genome reference assembly for variant calling. Variants were confirmed via Sanger sequencing using newly isolated gDNA extracted from the whole blood.

Specific PCR primers were designed using Primer3web (version 4.1.10; https://primer3.ut.ee)^57^ and Beacon Designer software (https://www.premierbiosoft.com/qOligo/Oligo.jsp?PID=1). Target specificity was confirmed *in silico* through BLAT alignment (sequence identity >95%, full coverage) and virtual PCR simulations within the UCSC Genome Browser; https://genome.ucsc.edu/)^58^. High-purity primers lacking secondary structures and with uniform melting temperatures (*Tm*), were synthesized by TIB Molbiol (Berlin, Germany).

### In silico analysis

The genomic sequence of *SPRED1* (ENSG00000118692) and its corresponding (1335 bp) CDS were obtained from the EMBL-EBI database (https://www.ebi.ac.uk)^59^. Clinical, phenotypic, and somatic distribution data associated with LS and cancer cross-referenced using the Leiden Open Variation Database LOVD (https://www.lovd.nl/)^60^ and IntOGen (https://www.intogen.org/search?gene=SPRED1)^55^ databases. Variant annotations were additionally compared with records from the historical ARUP *SPRED1* database, which was publicly available at the time of data collection.

The frequency, pathogenic prediction, and novelty of *SPRED1* variants were searched in ClinVar (https://www.ncbi.nlm.nih.gov/clinvar/)^61^, Varsome (https://varsome.com)^62^, Franklin (https://franklin.genoox.com/clinical-db/home)^63^, while somatic variants were obtained from the Catalogue of Somatic Mutations in Cancer (COSMIC) (https://cancer.sanger.ac.uk/cosmic)^15^. Population-level allele frequencies were determined by analyzing variant presence in the Genome Aggregation Database (gnomAD) (https://gnomAD.broadinstitute.org/)^64^, applying a two-tailed Fisher’s exact test with a significance threshold of p<0.05 for statistical validation and downstream cohort comparisons.

Position-specific evolutionary conservation across multiple species was quantified using PhyloP100 scores via Varsome and mapped through the ConSurf server (https://consurf.tau.ac.il/)^65^. To visualize the three-dimensional structural mapping of missense variations, the high-confidence predicted tertiary structure of SPRED1 was downloaded from the AlphaFold Protein Structure Database (https://alphafold.ebi.ac.uk/)^66^ in PDB format.

PyMOL (version 3.1; Schrodinger, LLC; https://www.pymol.org/) was used to generate the 3D structural templates and compute spatial interatomic distances at the neurofibromin interface. Variant location within functional domains was annotated using cBioPortal Mutation Mapper (https://www.cbioportal.org/mutation_mapper)^67^, integrated with UniProt database structural profiles (Entry:Q7Z699) (https://www.uniprot.org/uniprotkb/Q7Z699/entry)^68^. Serial Cloner software (version 2.6.1) was employed for *in silico* sequence alignment, restriction mapping and plasmid cloning design (http://serialbasics.free.fr/Serial_Cloner.html)^69^.

### Structural modeling and computational analysis of SPRED1 variants

Variants were retrieved from ClinVar and COSMIC and prioritized based on FATHMM-MKL pathogenicity score >0.7 or ClinVar annotation of “pathogenic” or “likely pathogenic”. Macromolecular analyses used the experimental crystal structure of the SPRED1–NF1–KRAS complex (PDB: 6V65) for the EVH1 domain and AlphaFold coordinates (UniProt: Q7Z699) for unresolved KBD and SPR regions. A FoldX (v5.0) pipeline was used to quantify structural stability (ΔΔG) and binding affinity. Ten independent runs were executed per variant. Residues within ≤6 Å of an interacting partner chain were considered interface residues. All calculations were performed under default parameters (298 K, 0.05 M ionic strength, pH 7.0).

### Cloning procedures

*SPRED1* gene was amplified by PCR from R777-E321 plasmid (Addgene, #70605) using custom-designed primers containing the human *SPRED1* open reading frame (ORF) [NM_152594.3]. The PCR primers were designed to flank the *SPRED1* sequence with SpeI (Thermo Fisher Scientific, Ref. FD1253) and NotI (Thermo Fisher Scientific, Ref. FD0594) restriction sites. The resulting PCR product was purified and cloned into the linearized EcoRV site of pNZY28 cloning vector (NZYTech, Ref. MB12101) to generate the intermediate construct pNZY28:*SPRED1*. The sequence was confirmed by Sanger sequencing. The SpeI/NotI restriction fragment containing the *SPRED1* insert was cut out and cloned into the pLVX-IRES-Hyg expression vector (Ref. 4064-5, Catalog N° 632185) previously digested with the same enzymes, generating the pLVX:*SPRED1* construct. *E. coli* DH5α was the host forroutine plasmid transformation and amplification. Bacterial cultures were grown at 37°C in LB liquid medium or agar plates supplemented with Ampicillin (100 µg/mL) to maintain plasmid selection.

For routine screening and sequencing, small-scale plasmid DNA isolation was performed using the GeneJet Plasmid Mini prep kit (Thermo Fisher Scientific, Ref. K0503). Large-scale plasmid preparations required for high-efficiency downstream transfections were isolated and purified utilizing the PureLink HiPure Plasmid Maxiprep kit (Invitrogen, Ref. K210017), following the manufacturer’s protocols. DNA fragments were extracted from agarose gels and purified with the NZYGelPure DNA Purification kit (NZYTech, Ref. MB011). Ligation reactions were performed overnight at room temperature using T4 DNA Ligase (Thermo Fisher Scientific, Ref. 15224041). DNA concentration and purity were determined by measuring absorbance at a wavelength of 260 nm using a NanoDrop™ spectrophotometer (Thermo Scientific™).

### Site-directed mutagenesis

The following *SPRED1* variants c.52C>T, c.184G>A, c.207+1G>A, c.305C>T, c.478G>T, c.634G>A, c.968C>A, c.986A>T, c.1048G>T, c.1089A>G and c.1318delC, were generated via PCR site-directed mutagenesis utilizing the intermediate plasmid pNZY28:*SPRED1* as the template using high-fidelity DNA polymerase (Platinum SuperFi II PCR Master Mix, Invitrogen, Ref. 12368010). PCR products were incubated with 1 μL *DpnI* (FastDigest *DpnI*; Thermo Fisher Scientific, Ref. FD1703) at 37°C.

### Lentiviral production and generation of stable cell lines

HEK293T cells (ATCC, Ref. CRL3216) were utilized for both lentiviral packaging and the subsequent generation of stable expression models. All cell lines were maintained in a humidified atmosphere at 37°C with 5% CO_2_. Cells were cultured in DMEM/F-12 GlutaMAX (Gibco®, Ref. A4192001) supplemented with 5% heat-inactivated fetal bovine serum (FBS; NeoBiotech, Ref. NB-03-0163). Lentiviruses were produced using a second-generation packaging system including the lentiviral expression vector (pLVX_IRES_Hyg and derivative constructs) and the packaging plasmids pVSV-G (Addgene, #138479) and psPAX2 (Addgene, #12260) using polyethylenimine. Viral supernatants were collected and filtered through 0.45 µm membranes, and used to transduce HEK293T cells. Stable integrants were selected with hygromycin B (150 µg/mL) for 10–14 days. All cultures were systematically screened and confirmed to be free of *Mycoplasma* contamination using a PCR-based detection kit (Vazyme, Cat. N°. D101).

### Western Blotting

The Western blot assays were conducted to analyze the protein levels of SPRED1, ERK1/2, AKT, p38, p53 and β-Actin, as well as the relative activation states via specific phosphorylation markers: ERK1/2 (phospho-ERK1/2 T202/Y204), AKT (phospho-AKT-T308, phospho-AKT-S473), p38 (phospho-p38 T180/Y182) and p53 (phospho-p53 S15). Stable cell lines were harvested and lysed in ice-cold RIPA lysis buffer (25 mM Tris-HCl, pH 7.6, 150 mM NaCl, 1% NP-40, 0.1% sodium deoxycholate, and SDS) supplemented with protease inhibitor (Thermo Fisher Scientific, Cat. A32955) and phosphatase inhibitor (Thermo Fisher Scientific, Cat. A32957) cocktails. Lysates were cleared by centrifugation at 12,000 x g for 15 min at 4°C, and supernatants were recollected. Total protein concentration was quantified using the Bradford assay.

The protein samples were separated on 10% SDS-PAGE gels (acrylamide/bis-acrylamide 29:1) for 60 minutes at 100 V in SDS-PAGE running buffer. Subsequently, the proteins were transferred onto nitrocellulose membranes with iBlot®2 NC Stacks (Invitrogen/ Thermo Fisher Scientific, Ref. IB23001). To prevent non-specific antibody binding, membranes were incubated in blocking buffer consist of Tris-buffered saline containing 0.1% Tween-20 (TBST) supplemented with 5% milk (Oxoid, Ref. LP0033B) for 1 hour at room temperature. Phospho-specific targets were detected in TBST containing 5% BSA (NZYTech Ref. MB47002). Membranes were incubated overnight at 4°C with the corresponding primary antibodies diluted in their respective blocking matrices: mouse anti- SPRED1 (1:1000; Ref. sc-393198, Santa Cruz Biotechnology), rabbit anti-total ERK1/2 (1:2000; Ref. 11257-1-AP, Proteintech), rabbit anti-phospho-ERK, p44/42 MAPK (ERK1/2) (1:1000; Ref. #9101 Cell Signaling Technology), rabbit anti-total AKT (1:1000; Ref. #9272, Cell Signaling Technology), rabbit anti-phospho-AKT S473 (1:1000; Ref. #9271, Cell Signaling Technology), rabbit anti-phospho-AKT T308 (1:1000; Ref. #9275, Cell Signaling Technology), mouse anti-total p38α MAPK14 (A1F7) (1:1000; Ref. sc-33688, Santa Cruz Biotechnology), rabbit anti-phospho-p38 MAPK (T180/Y182) (1:1000; Ref. #9211, Cell Signaling Technology), rabbit anti-total p53 (DO-1) (1:1000; Ref. sc-126, Santa Cruz Biotechnology), rabbit anti-phospho-p53 (S15) (1:1000; Ref. #9284 Cell Signaling Technology), mouse anti-β-Actin (1:4000; HRP-conjugated; Ref. 60008 Proteintech) and Rhodamine Anti-Actin hFAB (Ref. 12004164, Bio-Rad).

For chemiluminescent target development, membranes were incubated for 1 hour at room temperature with horseradish peroxidase (HRP)-conjugated secondary antibodies: goat anti-mouse IgG (1:10000; Invitrogen, Cat. #G21040) or goat anti-rabbit IgG (1:10000; Invitrogen, Cat. #G-21234) diluted in TBST-5% milk. The immunoreactive bands were visualized using the Pierce ECL Western Blotting Substrate (Thermo Fisher Scientific, Ref. 32106). For quantitative, fluorescence-based multiplexed detection, membranes were incubated for 1 hour at room temperature in the dark with Starbright™ fluorophore-conjugated secondary antibodies: anti-mouse (1:10000; Ref. 12004159, Bio-Rad) or anti-rabbit (1:10000; Ref. 12004162, Bio-Rad) diluted in TBST supplemented with 5% BSA, allowing direct target acquisition without exogenous chemical substrates.

Blot images were acquired and digitized using the Starbright secondary channel on the ChemiDoc™ MP Imaging System (Bio-Rad, Cat. #12003154). Densitometric quantification of band intensities was performed using Image Lab software (Bio-Rad), and normalized to the corresponding loading control. The number on independent biological replicates for each biomarker is specified in the Quantification and statistical analysis section. Data are expressed as mean ± standard error of the mean (SEM).

### RT-qPCR

Total RNA was isolated from HEK293T cells using the PureLink™ RNA Mini Kit (Cat. No. 12183025; Thermo Fisher Scientific, Waltham, MA, USA) according to the manufacturer’s protocols. Complementary DNA (cDNA) was synthesized using 1μg of total RNA template via the RevertAid First Strand cDNA Synthesis Kit (Cat. No. K1621) with random hexamer primers. qPCR assays were performed in 96-well microplates using the PowerTrack™ SYBR™ Green Master Mix (Thermo Fisher Scientific, Cat. No. A46012) on a QuantStudio™ 5 Real-Time PCR System to quantify *SPRED1* transcript levels. All reactions were performed in technical triplicates. Relative mRNA expression levels were calculated via the Pfaffl efficiency-corrected method, utilizing the *GAPDH* gene as the internal reference control. Specific primer sequences were as follows: *SPRED1*-Forward, 5’- AGACGGCGACTTCTGACAAC-3’; *SPRED1*-Reverse, 5’- ACAGTGACGCTGCTTAGTCC-3’; *GAPDH*-Forward, 5′-TGTTGCCATCAATGACCCCTT-3′; *GAPDH*-Reverse, 5′-CTCCACGACGTACTCAGCG-3′. Experiments were performed in three independent biological replicates (*n*=3) and data are presented as mean ± SEM.

### Real-time Cell Proliferation and Cytotoxicity Assays (IncuCyte®)

Stable HEK293T lines were seeded at 1 × 10l cells/well in 96-well plates (Falcon, REF 353072). Phase-contrast images were acquired every 6 h over 72 h using the IncuCyte® Live-Cell Analysis System (Sartorius). Cell proliferation was quantified using AI-based nuclei object counting and normalized to the initial time point (t = 0 h). Proliferation curves were generated over the entire acquisition period, and proliferation at 48 h were used for quantitative comparisons between experimental conditions. Assays were performed in three independent biological replicates (*n*=3), each containing triplicate technical wells per condition. Data are expressed as mean ± SEM.

### Quantification and statistical analysis

All statistical analyses, hypothesis testing, and analytical plot generation were executed utilizing GraphPad Prism software (version 10.0.2; GraphPad Software, San Diego, California, USA; www.graphpad.com) and R software (version 4.4.1; R Foundation for Statistical Computing, Vienna, Austria https://www.R-project.org/)^70^. Statistical analysis included one-way analysis of variance (ANOVA) followed by Dunnett’s post-hoc test to identify statistically significant deviations specifically against the WT reference control. To determine whether individual normalized variant ratios significantly deviated from a theoretical baseline reference value of 1.0 (such as the normalized AUC indices), a two-tailed one-sample *t*-test was systematically executed. Statistical significance thresholds across all tests were defined as a priori and categorized as follows: ****P<0.0001, ***P<0.001, **P<0.01, and *P<0.05. Values yielding P>0.05 were formally designated as non-significant (ns).

Correlation between signaling biomarkers and cell proliferation was assessed by Pearson correlation matrix analysis using a Multiple Variables table in GraphPad Prism, including p-ERK, p-AKT Ser473, and p-p38 as signaling variables. p-AKT Thr308 and p-p53 Ser15 were excluded from correlation analyses as neither showed statistically significant variation across the variant panel. All correlation analyses were performed on condition means (*n*=13 conditions) derived from the normalized ratio values used in the primary statistical comparisons.

Unsupervised hierarchical clustering and heatmap visualization were performed using ClustVis (https://biit.cs.ut.ee/clustvis/)^71^, a web-based tool for clustering and visualization of multivariate data. Signaling ratio values were row-centered and unit variance scaling was applied prior to clustering. Both row and column clustering were performed using Pearson correlation distance with average linkage.

All data are represented as mean ± SEM from at least three independent biological replicates. The number of independent biological replicates per assay included SPRED1 overexpression (*n*=5); ERK and p53 phosphorylation analyses, (*n*=5); AKT Ser 473, AKT Thr 308, and p38 phosphorylation analyses, (*n*=4); cell proliferation assays (*n*=3). Each biological replicate value represents the mean of three technical replicates performed within that experiment.

## Data Availability

All data produced in the present study are available upon reasonable request to the authors

## Ethics approval and consent to participate

This study was conducted in accordance with the ethical standards of the Declaration of Helsinki and its later amendments. Ethical approval was obtained from the Ethics Committee of the Bioethical Committee of the Institute of Biomedical Research of Salamanca (IBSAL) (Approval Code: PI 2022 07 1088 – GRA Project). Written informed consent was obtained from all participants (or their legal guardians in the case of minors or vulnerable populations) prior to their inclusion in the study. Participants were informed about the study’s purpose, procedures, potential risks, and their right to withdraw at any time without consequences. All data were anonymized to protect participant confidentiality.

## Consent for publication

Written informed consent for publication of clinical data was obtained from all participants or their legal guardians prior to inclusion in this study.

## Availability of data and materials

Due to privacy/ethical restrictions imposed by the Ethics Committee of the Bioethical Committee of the Institute of Biomedical Research of Salamanca (IBSAL), the raw data supporting this study regarding the patients cannot be made publicly available. However, anonymized data are available from the corresponding author upon reasonable request and with permission from the Ethics Committee. The rest of the data and materials regarding the experimental work are available upon request. Interested researchers should contact Prof. Jesus Lacal at to request access to the data.

## Funding

This study received support from the Alicia Koplowitz Foundation through the Research Grants program (Code: FAK21/001), Gerencia Regional de Salud de Castilla y León through the Research Grants program (Codes: GRS2548/A/22 and GRS2084/A/19), Sociedad Española de Endocrinología Pediátrica through the Research Grants program (Code: SEEP21/001), and University of Salamanca through the Research Grants program (Code: PIC2-2020-20), for laboratory equipment and personnel. Additional funding was provided by the Banco Santander (Research Mobility Grant 2022-24). The authors declare that the founding sources had no role in the study design, data analysis, or publication.

## Acknowledgments

We acknowledge the University Hospital of Salamanca for providing access to clinical facilities, Sacyl and IBSAL for their support in data collection, and University of Salamanca for administrative resources. We sincerely thank the lab technicians Sonia Andrés Recio, Fernando Díez Martín, and María León Martínez for their technical expertise. We also extend our gratitude to the students who were trained on this project: Eva Martínez, Jenny Rietdorf, Michelle Cerdan, Leonie Wagner, Beatrice Wagner, Flore Cuisin, Luna Misis and Tabea Hauer.

## Declaration of interests

The authors declare no competing interests.

## Author contributions

J.B.-F conducted all the experiments, performed the structural bioinformatics analysis, molecular modeling and carried out the data analysis. A.C analyzed the *in silico* data, while N.F performed the qPCR assays. Conceptualization, investigation, and writing—original draft preparation: J.L., J.B.-F., and P.P-M; writing, review and editing: J.L., J.B.-F., A.C., N.F., D.H.H-J, E.M-V., S.O., P.P-M and M.I-G; supervision: J.L.; funding acquisition: P.P.-M., M.I.-G., and J.L. All authors have read and agreed to the published version of the manuscript.

## Supplementary Tables and Figures

**Supplementary Table S1. *SPRED1* variants annotated in ClinVar.** As of July 25, 2024, ClinVar has annotated up to 846 variants. In yellow, 112 predicted pathogenic variants in *SPRED1* were examined for their distribution.

Supplementary Table S2. Comprehensive biophysical, geometric, and computational energetic profiling of *SPRED1* variants within the EVH1 domain interface. Data represent mean ± SEM derived from ten independent FoldX simulation runs (*n* = 10). Positive ΔΔG values denote monomer destabilization or decreased complex binding affinity, whereas negative values indicate relative stabilization compared to the WT baseline. Geometric properties of the docking interface were quantified via Buried Surface Area (BSA) calculations and the total number of interatomic contacts within a ≤ 6 Å threshold distance. Abbreviations: aa, amino acids; IE, Interaction Energy; BSA, Buried Surface Area; PTC-induced, structural metrics following simulated downstream sequence cleavage; N/A, not applicable for this variant class.

**Supplementary Figure 1. Effects of *SPRED1* variants on transcript and SPRED1 endogenous levels in c.52C>T variant. (a)** Relative *SPRED1* mRNA expression levels analyzed by RT-qPCR in stable HEK293T cell lines expressing WT or mutant constructs. Transcript levels were normalized to the WT reference control group (set at 1.0) and computed utilizing the efficiency-corrected Pfaffl method with *GAPDH* as the internal reference gene. Statistical significance was determined by a one-way ANOVA followed by Dunnett’s post-hoc test for multiple comparisons against the WT control ***p < 0.001, ****p < 0.0001. Data represent mean ± SEM from three independent biological replicates (*n* = 3). High transcript abundance confirms successful lentiviral transcription despite the lack of protein detection for the c.52C>T and c.207+1G>A variants. **(b)** Immunoblot characterization of endogenous SPRED1 protein expression levels in cells expressing the c.52C>T variant. Representative samples were resolved independently via WB alongside parental and MOCK controls to corroborate the apparent compensatory upregulation or stabilization of a protein band corresponding to the molecular weight of WT endogenous SPRED1, suggesting an alteration in proteasomal turnover or a regulatory homeostatic feedback loop induced by the truncated peptide architecture.

**Supplementary Figure 2. (a)** Area under the curve (AUC) analysis of proliferation kinetics. Total area under the proliferation curve (AI nuclei object count fold-change vs. time) calculated over the full 72-hour kinetic window for all stable cell lines. AUC values are normalized to WT (set as 1.00) and represent the mean of three independent biological replicates (*n*=3). The c.478G>T variant showed the lowest normalized AUC (0.584 relative to WT), consistent with its 48h proliferation value (0.469 relative to WT), confirming that progressive kinetic deceleration accumulates into substantially reduced overall proliferative output. AUC analysis confirms that proliferative differences observed at 48h reflect sustained growth trajectories rather than transient responses at a single time point. **(b)** Pearson correlation matrix of signaling biomarkers and cell proliferation showing pairwise correlation coefficients (r) between phospho-ERK1/2 (T202/Y204), phospho-AKT Ser473, phospho-p38 (T180/Y182), and cell proliferation at 48h across the thirteen conditions analyzed (WT and eleven SPRED1 variants plus MOCK). Values represent Pearson r coefficients calculated from condition means. Color scale ranges from −1 (blue, negative correlation) to +1 (red, positive correlation). None of the associations between individual signaling biomarkers and proliferation reached statistical significance (all p>0.05; *n*=13 conditions). Phospho-AKT Thr308 and phospho-p53 Ser15 were excluded from the analysis as neither showed statistically significant variation across the variant panel. Analysis performed using GraphPad Prism 10 (Multiple Variables table, Pearson method, two-tailed). **(c)** Unsupervised hierarchical clustering of multi-pathway signaling and proliferation data. Heatmap showing unsupervised hierarchical clustering of normalized signaling ratio values (phospho-ERK1/2, phospho-AKT Ser473, phospho-p38, and cell proliferation at 48h) across all thirteen conditions. Values were row-centered and unit variance scaling was applied prior to clustering. Both row and column clustering were performed using Pearson correlation distance with average linkage. Color scale represents scaled expression values ranging from −1 (blue, below mean) to +1 (red, above mean).

## Notes

### Competing Interest Statement

The authors have declared no competing interest.

### Author Declarations

The Bioethical Committee of the Institute of Biomedical Research of Salamanca (IBSAL) gave ethical approval for this work. Written informed consent for publication of identifiable images (including facial features) and clinical data was obtained from all participants (or their legal guardians if under 18 years old), except for one family who declined consent for facial images. Consequently, these images have been excluded from publication. A copy of the consent forms is available for review by the journal's editorial team upon request.

## References

1. Montero-Bullón, J. F., González-Velasco, Ó., Isidoro-García, M. & Lacal, J. Integrated in silico MS-based phosphoproteomics and network enrichment analysis of RASopathy proteins. Orphanet J. Rare Dis. 16, 1–17 (2021).

2. Brems, H. et al. Germline loss-of-function mutations in SPRED1 cause a neurofibromatosis 1-like phenotype. Nat. Genet. 39, 1120–1126 (2007).

3. Stowe, I. B. et al. A shared molecular mechanism underlies the human rasopathies Legius syndrome and Neurofibromatosis-1. Genes Dev. 26, 1421–1426 (2012).

4. Messiaen, L. et al. Clinical and mutational spectrum of neurofibromatosis type 1-like syndrome. JAMA - J. Am. Med. Assoc. 302, 2111–2118 (2009).

5. Denayer, E. et al. Observations on intelligence and behavior in 15 patients with Legius syndrome. Am. J. Med. Genet. Part C Semin. Med. Genet. 157, 123–128 (2011).

6. Giugliano, T. et al. Clinical and genetic findings in children with neurofibromatosis type 1, legius syndrome, and other related neurocutaneous disorders. Genes (Basel). 10, (2019).

7. Pasmant, E. et al. Neurofibromatosis type 1 molecular diagnosis: What can NGS do for you when you have a large gene with loss of function mutations? Eur. J. Hum. Genet. 23, 596–601 (2015).

8. Evans, D. G. R. et al. Comprehensive RNA Analysis of the NF1 Gene in Classically Affected NF1 Affected Individuals Meeting NIH Criteria has High Sensitivity and Mutation Negative Testing is Reassuring in Isolated Cases With Pigmentary Features Only. eBioMedicine 7, 212–220 (2016).

9. Fowlkes, J. L., Thrailkill, K. M. & Bunn, R. C. RASopathies: The musculoskeletal consequences and their etiology and pathogenesis. Bone 152, 116060 (2021).

10. Xi, W. et al. Clinical and genetic analysis of three children with Legius syndrome due to variants of SPRED1 gene. Chinese J. Genet. J. Genet. 41, 941–946 (2024).

11. Pasmant, E. et al. SPRED1 disorder and predisposition to leukemia in children. Blood 114, 1131 (2009).

12. Lorenzo, C. & McCormick, F. SPRED proteins and their roles in signal transduction, development, and malignancy. Genes Dev. 34, 1410–1421 (2020).

13. Pasmant, E. et al. SPRED1, a RAS MAPK pathway inhibitor that causes Legius syndrome, is a tumour suppressor downregulated in paediatric acute myeloblastic leukaemia. Oncogene 34, 631–638 (2015).

14. Spencer, E. et al. Identification of SPRED1 deletions using RT-PCR, multiplex ligation-dependent probe amplification and quantitative PCR. Am. J. Med. Genet. Part A 155, 1352–1359 (2011).

15. Sondka, Z. et al. COSMIC: a curated database of somatic variants and clinical data for cancer. Nucleic Acids Res. 52, D1210–D1217 (2024).

16. Pasmant, E. et al. SPRED1 germline mutations caused a neurofibromatosis type 1 overlapping phenotype. J. Med. Genet. 46, 425–430 (2009).

17. Spurlock, G. et al. SPRED1 mutations (Legius syndrome): Another clinically useful genotype for dissecting the neurofibromatosis type 1 phenotype. J. Med. Genet. 46, 431–437 (2009).

18. Brems, H. et al. Review and update of SPRED1 mutations causing legius syndrome. Hum. Mutat. 33, 1538–1546 (2012).

19. Cemeli Cano, M., et al. Un nuevo síndrome neurocutáneo: síndrome de Legius. A propósito de un caso. Rev. Neurol. 59, 209 (2014).

20. Bianchi, M. et al. Legius Syndrome: two novel mutations in the SPRED1 gene. Hum. Genome Var. 2, 2–4 (2015).

21. Kimura, R., Yoshida, Y., Maruoka, R., Kosaki, K. & Yamamoto, O. Legius syndrome: A case report. J. Dermatol. 44, 459–460 (2017).

22. Ablain, J. et al. Human tumor genomics and zebrafish modeling identify SPRED1 loss as a driver of mucosal melanoma. Science (80-.). 362, 1055–1060 (2018).

23. Liu, W., Fang, S. & Zuo, G. A study on the expression of SPRED1 and PBRM1 (Baf180) and their clinical significances in patients with gastric cancer. Clin. Lab. 6, 2117–2124 (2020).

24. Wang, J. et al. mir182 activates the Ras–MEK–ERK pathway in human oral cavity squamous cell carcinoma by suppressing RASA1 and SPRED1. Onco. Targets. Ther. 10, 667–679 (2017).

25. Jiang, C. F. et al. Estrogen-induced miR-196a elevation promotes tumor growth and metastasis via targeting SPRED1 in breast cancer. Mol. Cancer 17, (2018).

26. Yoshida, T. et al. Spreds, inhibitors of the Ras/ERK signal transduction, are dysregulated in human hepatocellular carcinoma and linked to the malignant phenotype of tumors. Oncogene 25, 6056–6066 (2006).

27. Zhang, R. et al. SPRED1 Is Downregulated and a Prognostic Biomarker in Adult Acute Myeloid Leukemia. Front. Oncol. 10, 1–13 (2020).

28. Ullrich, M. et al. Identification of SPRED2 (Sprouty-related protein with EVH1 domain 2) as a negative regulator of the hypothalamic-pituitary-adrenal axis. J. Biol. Chem. 286, 9477–9488 (2011).

29. Silva, J. M., Canche, L., Cheng, A., Young, L. C. & McCormick, F. NF1 and SPRED1/2 cooperate through RAS-MAPK-independent functions. Proc. Natl. Acad. Sci. U. S. A. 123, e2535319123 (2026).

30. Siljamäki, E. & Abankwa, D. SPRED1 Interferes with K-ras but Not H-ras Membrane Anchorage and Signaling. Mol. Cell. Biol. 36, 2612–2625 (2016).

31. Dunzendorfer-Matt, T., Mercado, E. L., Maly, K., McCormick, F. & Scheffzek, K. The neurofibromin recruitment factor Spred1 binds to the GAP related domain without affecting Ras inactivation. Proc. Natl. Acad. Sci. U. S. A. 113, 7497–7502 (2016).

32. Hirata, Y. et al. Interaction between a domain of the negative regulator of the ras-ERK pathway, SPRED1 protein, and the GTPase-activating protein-related domain of neurofibromin is implicated in legius syndrome and neurofibromatosis type 1. J. Biol. Chem. 291, 3124–3134 (2016).

33. Chaker-Margot, M. et al. Structural basis of activation of the tumor suppressor protein neurofibromin. Mol. Cell 82, 1288–1296.e5 (2022).

34. Yan, W. et al. Structural Insights into the SPRED1-Neurofibromin-KRAS Complex and Disruption of SPRED1-Neurofibromin Interaction by Oncogenic EGFR. Cell Rep. 32, 107909 (2020).

35. Wakioka, T. et al. Spred is a sprouty-related suppressor of Ras signalling. Nature 412, 647–651 (2001).

36. Hirata, Y. et al. Legius syndrome mutations in the Ras-regulator SPRED1 abolish its membrane localization and potentially cause neurodegeneration. J. Biol. Chem. 300, 107969 (2024).

37. Sumner, K. et al. The SPRED1 variants repository for legius syndrome. G3 Genes, Genomes, Genet. 1, 451–456 (2011).

38. Führer, S., Tollinger, M. & Dunzendorfer-Matt, T. Pathogenic Mutations Associated with Legius Syndrome Modify the Spred1 Surface and Are Involved in Direct Binding to the Ras Inactivator Neurofibromin. J. Mol. Biol. 431, 3889–3899 (2019).

39. Kato, R. et al. Molecular cloning of mammalian Spred-3 which suppresses tyrosine kinase-mediated Erk activation. Biochem. Biophys. Res. Commun. 302, 767–772 (2003).

40. Nonami, A. et al. Spred-1 negatively regulates interleukin-3-mediated ERK/mitogen-activated protein (MAP) kinase activation in hematopoietic cells. J. Biol. Chem. 279, 52543–52551 (2004).

41. Führer, S. et al. NMR resonance assignments of the EVH1 domain of neurofibromin’s recruitment factor Spred1. Biomol. NMR Assign. 11, 305–308 (2017).

42. Butland, S. L. et al. The palmitoyl acyltransferase HIP14 shares a high proportion of interactors with huntingtin: Implications for a role in the pathogenesis of Huntington’s disease. Hum. Mol. Genet. 23, 4142–4160 (2014).

43. Nonami, A. et al. The Sprouty-related protein, Spred-1, localizes in a lipid raft/caveola and inhibits ERK activation in collaboration with caveolin-1. Genes to Cells 10, 887–895 (2005).

44. Suzuki, M., Morita, R., Hirata, Y., Shichita, T. & Yoshimura, A. Spred1, a Suppressor of the Ras–ERK Pathway, Negatively Regulates Expansion and Function of Group 2 Innate Lymphoid Cells. J. Immunol. 195, 1273–1281 (2015).

45. Wei, L. et al. Oxidative Stress-mediated Sprouty-related Protein with an EVH1 Domain 1 Down-regulation Contributes to Resisting Oxidative Injury in Microglia. Neuroscience 526, 13–20 (2023).

46. Li, D., Jackson, R. A., Yusoff, P. & Guy, G. R. Direct association of sprouty-related protein with an EVH1 domain (SPRED) 1 or SPRED2 with DYRK1A modifies substrate/kinase interactions. J. Biol. Chem. 285, 35374–35385 (2010).

47. Douben, H. et al. Functional Assays Combined with Pre-mRNA-Splicing Analysis Improve Variant Classification and Diagnostics for Individuals with Neurofibromatosis Type 1 and Legius Syndrome. Hum. Mutat. 2023, 9628049 (2023).

48. Hoang, D. H. et al. MicroRNA networks in FLT3-ITD acute myeloid leukemia. Proc. Natl. Acad. Sci. U. S. A. 119, e2112482119 (2022).

49. King, J. A. J. et al. Distinct requirements for the Sprouty domain for functional activity of Spred proteins. Biochem. J. 388, 445–454 (2005).

50. Johne, C. et al. Spred1 and TESK1 - Two new interaction partners of the kinase MARKK/TAO1 that link the microtubule and actin cytoskeleton. Mol. Biol. Cell 19, 1391–1403 (2008).

51. Chelleri, C. et al. Novel causative variants in Legius syndrome: SPRED1 Genotype spectrum expansion. Am. J. Med. Genet. Part A 1–6 (2024) doi:10.1002/ajmg.a.63824.

52. Han, J. Y. & Park, J. Paternally Inherited Noonan Syndrome Caused by a PTPN11 Variant May Exhibit Mild Symptoms: A Case Report and Literature Review. Genes (Basel). 15, 445 (2024).

53. Athota, J. P. et al. Molecular and clinical studies in 107 Noonan syndrome affected individuals with PTPN11 mutations. BMC Med. Genet. 21, 50 (2020).

54. der Auweraer, S. Van et al. Generation and characterization of four iPSC and isogenic gene-corrected lines from Legius syndrome patients. Stem Cell Res. 94, 104026 (2026).

55. Gonzalez-Perez, A. et al. IntOGen-mutations identifies cancer drivers across tumor types. Nat. Methods 10, 1081–1084 (2013).

56. Raschka, S., Patterson, J. & Nolet, C. Machine learning in python: Main developments and technology trends in data science, machine learning, and artificial intelligence. Information (Switzerland) vol. 11 (2020).

57. Untergasser, A. et al. Primer3-new capabilities and interfaces. Nucleic Acids Res. 40, e115 (2012).

58. Navarro Gonzalez, J., et al. The UCSC genome browser database: 2021 update. Nucleic Acids Res. 49, D1046–D1057 (2021).

59. Emboss. EMBOSS Needle Pairwise Sequence Alignment. vol. Mai 2012 http://www.ebi.ac.uk/Tools/psa/emboss_needle/#.WHTuk7j4Yqg.mendeley%5Cnhttp://www.ebi.ac.uk/Tools/psa/emboss_needle/ (2013).

60. Fokkema, I. F. A. C. et al. LOVD v.2.0: The next generation in gene variant databases. Hum. Mutat. 32, 557–563 (2011).

61. Landrum MJ, Lee JM, Benson M, Brown GR, Chao C, Chitipiralla S, Gu B, Hart J, Hoffman D, Jang W, Karapetyan K, Katz K, Liu C, Maddipatla Z, Malheiro A, McDaniel K, Ovetsky M , Riley G, Zhou G, Holmes JB, Kattman BL, M. D. ClinVar. https://www.ncbi.nlm.nih.gov/clinvar/ (2018).

62. Kopanos, C. et al. VarSome: the human genomic variant search engine. Bioinformatics 35, 1978–1980 (2019).

63. Genoox, F. Franklin by Genoox. https://franklin.genoox.com/clinical-db/home (2021).

64. Gudmundsson, S. et al. Variant interpretation using population databases: Lessons from gnomAD. Hum. Mutat. 43, 1012–1030 (2022).

65. Ashkenazy, H. et al. ConSurf 2016: an improved methodology to estimate and visualize evolutionary conservation in macromolecules. Nucleic Acids Res. 44, W344–W350 (2016).

66. Varadi, M. et al. AlphaFold Protein Structure Database: Massively expanding the structural coverage of protein-sequence space with high-accuracy models. Nucleic Acids Res. 50, D439–D444 (2022).

67. Cerami, E. et al. The cBio Cancer Genomics Portal: An open platform for exploring multidimensional cancer genomics data. Cancer Discov. 2, 401–404 (2012).

68. Bateman, A. et al. UniProt: the Universal Protein Knowledgebase in 2025. Nucleic Acids Res. 53, D609–D617 (2025).

69. Michael Goodson, Philippe Benaroch, Helen Schreiner, Heiko Flammann, Cariappa Annaiah, Tatiana Nedvetskaya, Leopoldo Palma, Jolita Seckute, ATSbio, Benjamin Dickins, Juliette Azimzadeh, Paris Margaritis, Javier Irazoqui, Hideshi Yagi, M. P. y J. L. Clonador en serie. http://serialbasics.free.fr/Serial_Cloner.html (2013).

70. R Studio Team. A language and environment for statistical computing. R Found. Stat. Comput. 3, https://www.R-project.org (2021).

71. Metsalu, T. & Vilo, J. ClustVis: A web tool for visualizing clustering of multivariate data using Principal Component Analysis and heatmap. Nucleic Acids Res. 43, W566–W570 (2015).

