## Supplementary figures and images for "*SPRED1* variants reveal differential impacts on signaling dynamics"

### Figure S1

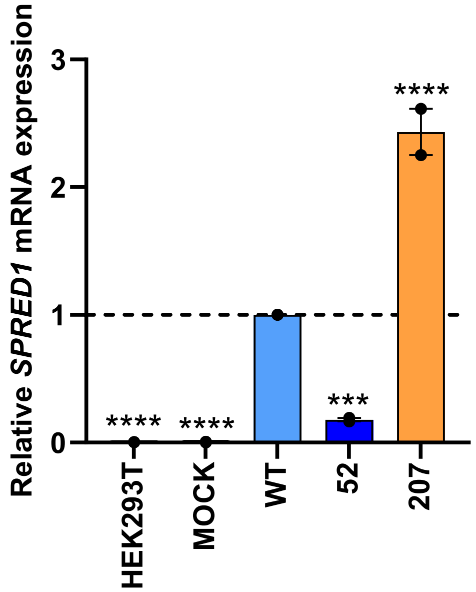

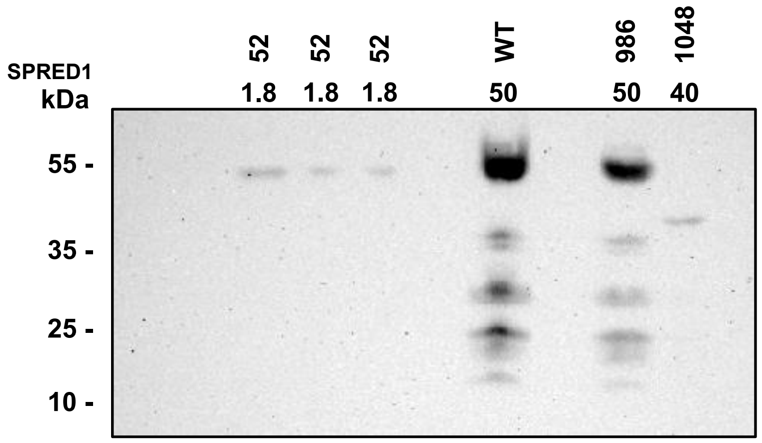

### Figure S2

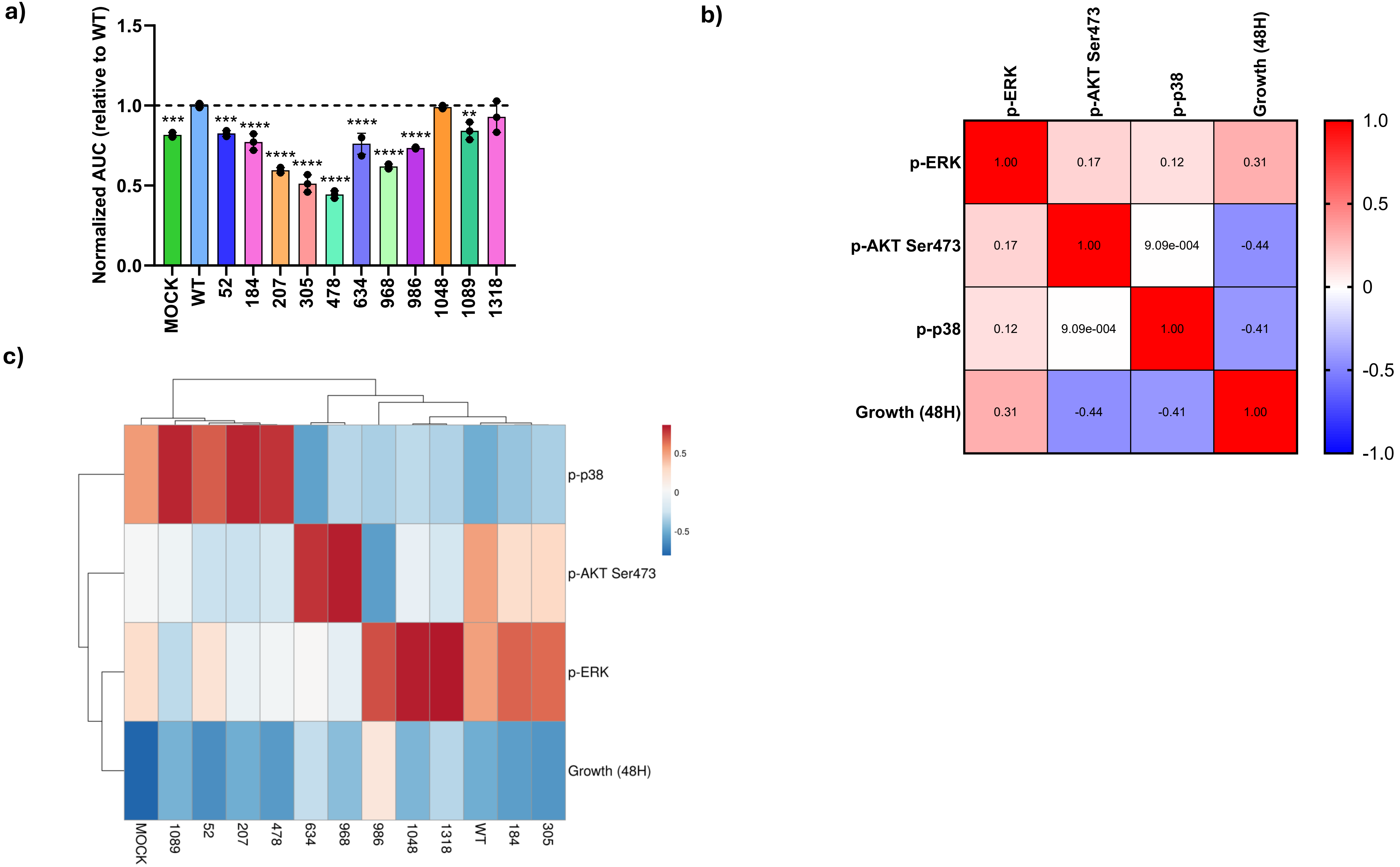
