## Supplementary material for "*SPRED1* variants reveal differential impacts on signaling dynamics": Table S2

| **Variant ID** | **Modification** | **Lenght**  **(aa)** | **Total Energy**  **(kcal/mol)** | **Mean Stability per residue (ST/aa)** | **ΔΔG folding**  **(kcal/mol)** | **ΔΔG_binding**  **(kcal/mol)** | **IE (kcal/mol)** |
| --- | --- | --- | --- | --- | --- | --- | --- |
| **WT** | Baseliine control | 113 | -65.611 | -0.580 | 0.000 | 0.000 | -17.774 |
| **G62R** | Missense substitution | 113 | N/A | N/A | +6.39 ± 2.20 | 0.000 ± 0.020 | N/A |
| **T102M** | Missense substitution | 113 | N/A | N/A | -1.95 ± 0.01 | -0.58 ± 0.30 | N/A |
